# An Explainable Multi-Modal Neural Network Architecture for Predicting Epilepsy Comorbidities Based on Administrative Claims Data

**DOI:** 10.1101/2020.11.07.20227454

**Authors:** Thomas Linden, Johann de Jong, Chao Lu, Victor Kiri, Kathrin Haeffs, Holger Fröhlich

## Abstract

Epilepsy is a complex brain disorder characterized by repetitive seizure events. Epilepsy patients often suffer from various and severe physical and psychological co-morbidities (e.g. anxiety, migraine, stroke, etc.). While general comorbidity prevalences and incidences can be estimated from epidemiological data, such an approach does not take into account that actual patient specific risks can depend on various individual factors, including medication. This motivates to develop a machine learning approach for predicting risks of future comorbidities for the individual epilepsy patient.

In this work we use inpatient and outpatient administrative health claims data of around 19,500 US epilepsy patients. We suggest a dedicated multi-modal neural network architecture (Deep personalized **LO**ngitudinal convolutional **RI**sk model - DeepLORI) to predict the time dependent risk of six common comorbidities of epilepsy patients. We demonstrate superior performance of DeepLORI in a comparison with several existing methods Moreover, we show that DeepLORI based predictions can be interpreted on the level of individual patients. Using a game theoretic approach, we identify relevant features in DeepLORI models and demonstrate that model predictions are explainable in the light of existing knowledge about the disease. Finally, we validate the model on independent data from around 97,000 patients, showing good generalization and stable prediction performance over time.

## 2 Introduction

Epilepsy is a complex, life threatening brain disorder characterized by repetitive seizure events. Epilepsy patients often suffer from various and severe physical and psychological comorbidities, such as overweight and obesity, anxiety, migraine, bipolar disorder and cardiovascular diseases (Seidenberg, Pulsipher and Hermann, 2009; Ottman *et al*., 2011; Keezer, Sisodiya and Sander, 2016). Some comorbidities confer a poor disease prognosis, because they complicate pharmacological treatment owing to possible drug-drug interactions and adverse events (Verrotti and Mazzocchetti, 2016). The actual development of comorbidities is dependent on patient specific factors and may be modulated by anti-epileptic drug (AED) treatment (Zaccara, 2009). Early identification and treatment of comorbidities has thus been identified as highly relevant to improve the quality of life of epilepsy patients (Verrotti and Mazzocchetti, 2016). However, there is a high subject to subject variability. Methods from the field of Artificial Intelligence (AI) and more specifically machine learning (ML) have the potential to predict comorbidity risks on an individual subject basis, hence fulfilling one of the promises of a more individualized patient care in the sense of precision medicine. More specifically, ML based approaches can be used to aid disease prevention by predicting the time dependent risk of an individual epilepsy patient to develop several common comorbidities in the future, such as (1) anxiety, (2) bipolar disorder & schizophrenia, (3) diabetes type 2, (4) migraine, (5) overweight & obesity, and (6) stroke & ischemic attacks.

Machine learning models to predict individualized comorbidity risks of diseases different from epilepsy have recently been published e.g. in (Dworzynski *et al*., 2020) and (Noh *et al*., 2020) using clinical routine data from the Danish national registry and hospital electronic health records, respecitvely. For epilepsy, (Glauser *et al*., 2020) proposed an ML model for psychatric comorbidities based on survey data from 122 patients. In our earlier work (Gerlach, Lu and Fröhlich, 2017) we proposed an ML model (Random Survival Forests) using US administrative health claims data from ∼10,000 epilepsy patients to predict several major comorbidities (anxiety, bipolar disorder & schizophrenia, diabetes type 2, migraine, overweight & obesity, stroke & ischemic attacks) of epilepsy patients.

Administrative health claims data have generally been shown useful for developing ML models in the epilepsy field. For example (An *et al*., 2018) used claims data of more than 1.3 Million epilepsy patients to predict antiepileptic drug resistance. Examples from other disease areas include prediction of Alzheimer’s Disease (Park *et al*., 2020), osteoporotic hip fractures (Engels *et al*., 2020), and heart failure (Desai *et al*., 2020). The opportunities of healthcare claims data for ML based modeling have further been discussed in (Fröhlich *et al*., 2018; Miotto *et al*., 2018; Xiao, Choi and Sun, 2018; Thesmar *et al*., 2019; Kwak and Hui, 2020).

In our earlier work we demonstrated the possibility to augment claims data with biomedical background knowledge, hence enabling the interpretation of machine learning models down to the level of disease associated biological processes (Gerlach, Lu and Fröhlich, 2017). The particular novelty of the present work is a dedicated multi-modal neural network architecture for administrative claims data, which we call **Deep** personalized **LO**ngitudinal convolutional **RI**sk model (DeepLORI). We show that DeepLORI more accurately predicts the time dependent risk for six common comorbidities on the level of individual patients than several competing methods, including our own previously proposed model. Using a game theoretic approach based on Shapley Additive Explanations (Lundberg and Lee, 2017) we show that DeepLORI models are explainable, also on the level of predictions for individual patients.

## 3 Data

### 3.1 Claims Based Electronic Health Records

US commercial inpatient and outpatient data were obtained from the IBM® MarketScan® Truven Health databases. The Commercial Claims and Encounters database within MarketScan® is a nationally representative collection of de-identified patient-specific inpatient, outpatient, and pharmaceutical claims from more than 200 insurance carriers and large, self-insuring companies. All dates and timestamps were transformed from a daily to a monthly scale (1 *month* := 30 *days*) for a more robust representation. The data generally comprises demographic (age, gender) and regional information (major metropolitan area), days in hospital, health insurance plan, as well as time dependent diagnosis codes and prescriptions (plus prescription duration and quantity).

We used two cohorts, (1) the *original data* covering years 2011-2015 for model-training and –evaluation within a nested cross-validation scheme, and (2) the *external validation data* covering years 2008-2018 to validate the models trained on the “original data”. In agreement to our earlier publication (Gerlach, Lu and Fröhlich, 2017) and common practice at UCB, epilepsy patients in the original data were identified matching at least one of the following criteria:

1. an occurrence of ≥2 ICD-9-CM codes of 345.xx (i.e. *epilepsy*, except 345.3 – *grand mal status*) among separate medical encounters (separate dates in any care venue)
2. an occurrence of ≥1 ICD-9-CM code of 345.xx (except for 345.3) AND ≥1 ICD-9-CM code of 780.39 (*convulsions*) among separate medical encounters
3. an occurrence of 1 ICD-9-CM code of 345.xx (except for 345.3) AND code(s) for AED prescription at least a day after the 345.xx code
4. an occurrence of ≥ 2 ICD codes of 780.39 among separate medical encounters AND code(s) for AED treatment. The code(s) for the AED treatment should occur at least a day after the second 780.39 irrespective of the presence or absence of an AED code after the first 780.39 code
5. Individuals with ICD-9-CM code 345.3 will be required to have an occurrence of ≥2 ICD-9-CM codes of 345.3 separated by at least 30 days, or an occurrence of the 345.3 code and ≥1 ICD-9-CM code 780.39 separated by at least 30 days, or ≥1 ICD-9-CM code 345.3 and ≥1 ICD-9-CM code 345.xx encounters on separate days

The index date for each patient was defined as the time point of the first epilepsy diagnosis, and for definitions requiring at least 2 ICD-9-CM codes the **first** diagnosis code was the index date. The data was further filtered by requiring for each patient a) at least 365 days of medical history before, and 365 days follow up after the index date; b) age between 18 and 65; c) any AED treatment during observation period. Altogether this yielded 7,430,840 records from 19,510 patients. For part of these patients diagnoses after index date were coded in ICD10, which we mapped to ICD-9-CM via the Thomas Reuters™ public web resource^2^ and manual curation.

Note that in medical practice, confirmation of the final diagnosis “epilepsy” can be complicated and often requires a number of visits. Moreover, reporting of a dedicated diagnosis within our data does not necessarily correspond to the actual time point of the medical condition within the patient. To capture this uncertainty we defined a three month time interval starting from the index date as "epilepsy diagnosis period”. That means the actual medical history of each patient after application of the above mentioned filter criteria was 365 + 91 days, i.e. 456 days.

Diagnosis codes after 1^st^ Oct 2015 were provided as ICD-10-CM codes. Accordingly, the following modified inclusion criteria were applied to select epilepsy patients in the external validation data (covering 2008-2018):

1. The presence of at least 1 ICD-9-CM of 345.xx or ICD-10-CM of G40.xx (*epilepsy*)
2. The presence of at least 2 ICD-9-CM of 780.39 or ICD-10-CM of R56.9 (*convulsions*) within one year.

After applying the same filter criteria as for the original data this resulted in 112,755 patients. Within those patients we prefiltered diagnosis codes and substances observed in ≤ 10 patients or with a frequency ≤ 1%.

One of the main issues with claims data is that one and the same diagnosis may be coded with different ICD codes. Moreover, observations related to one specific ICD9/10 code could be rare. To address these issues, we mapped all ICD-9-CM codes to PheWAS terms, which describe a higher level aggregate of several ICD codes (Carroll, Bastarache and Denny, 2014). In addition, a mapping to MeSH (Rogers, 1963) was performed to allow for integration with other data sources (see Methods).

### 3.2 Definition of Focused Comorbidities and Compilation of Training Data

Based on the medical literature as well as observed frequency in our data we focused on six common comorbidities of epilepsy patients: (1) Anxiety, (2) Bipolar and Schizophrenia, (3) Diabetes type 2, (4) Migraine, (5) Overweight and Obesity, (6) Stroke and Ischemic Attack. These comorbidities were defined according to a set of PheWAS codes provided in the Supplements (Table S1).

The number of patients with these comorbidities being diagnosed at least 6 months after the epilepsy diagnosis period differs widely across comorbidities, see **Table 1**. We would like to highlight that our data is in principle *right censored*, i.e. the diagnosis of a specific comorbiditiy might happen after the end of the period covered by our training data. Moreover, a significant proportion of those individuals where a diagnosis of a specific comoridity is observed (i.e. incident cases) have already been diagnosed with at least one of the other 5 comorbidities during their medical history, see **Table 1**. Note that for training a machine learning model to predict a specific comorbidity we should not have an observation of the same comorbidity in the medical history of any of the training samples. For this reason the number of patients in the training data is different per comorbidity, and we developed separate machine learning models for each comorbidity.

**Table 1:**
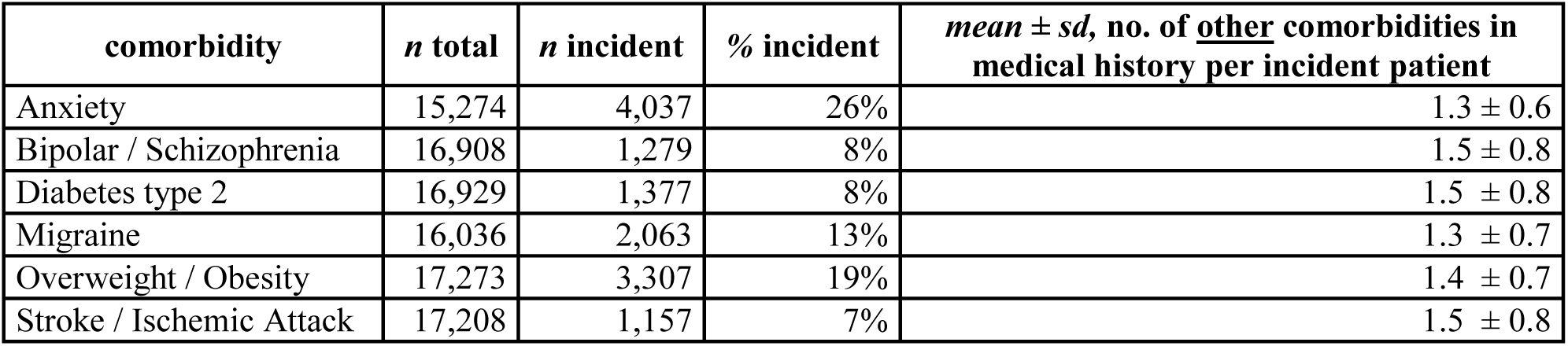
Proportion of incident patients by comorbidity.

Each diagnosis and prescription in our data has an associated time stamp. Due to the fact that the appearance of a record in our data does not necessarily correspond to the observation of the actual medical condition, each time stamp was mapped to a monthly (= 30 days time interval) resolution.

## 4 Methods

### 4.1 Proposed Model: DeepLORI

#### 4.1.1 DeepLORI Architecture

As highlighted before, our aim was to develop separate machine learning models for each of 6 typical comorbidities of epilepsy patients. Each of these models aims for predicting the time dependent risk of an individual to be diagnosed with one specific comorbidity.

We came up with a dedicated neural network model for our purposes, which we call **Deep** personalized **LO**ngitudinal convolution **RI**sk model (DeepLORI). We start by explaining the principle architecture of DeepLORI. In agreement to our former work, one of the key ideas is that claims data has an inherent hierarchical structure (Gerlach, Lu and Fröhlich, 2017): The data initially contains three major types of features: 1) prescribed substance codes, 2) diagnoses codes (mapped to PheWAS terms, see above) and 3) general demographic information, such as age, gender and major metropolitan area information. Monthly reported prescriptions and diagnoses can typically be represented via a one-hot vector encoding. However, individual substance and diagnose codes are typically rather sparsely observed over time, which can potentially lead to challenges for a machine learning algorithm to find regularities.

Based on this consideration our idea was to use additional background knowledge available in databases to impose further hierarchical sub-structure: For example, each prescribed substance may have one or several known targets, and it can have a number of side effects reported in clinical studies. Diagnoses have associated symptoms, and in some cases biomarkers may exist. Based on this background information each domain of features (e.g. diagnosis) can be further associated to several sub-domains (e.g. biomarkers, impaired biological pathways, etc.). Sub-domain features can subsequently be represented via a one-hot vector encoding. **Figure 1** and **Table 2** provide an overview about the domains and according sub-domains we defined in our data. More details about our previously published approach to augment claims data with biomedical knowledge can be found in the Supplements.

**Table 2:**
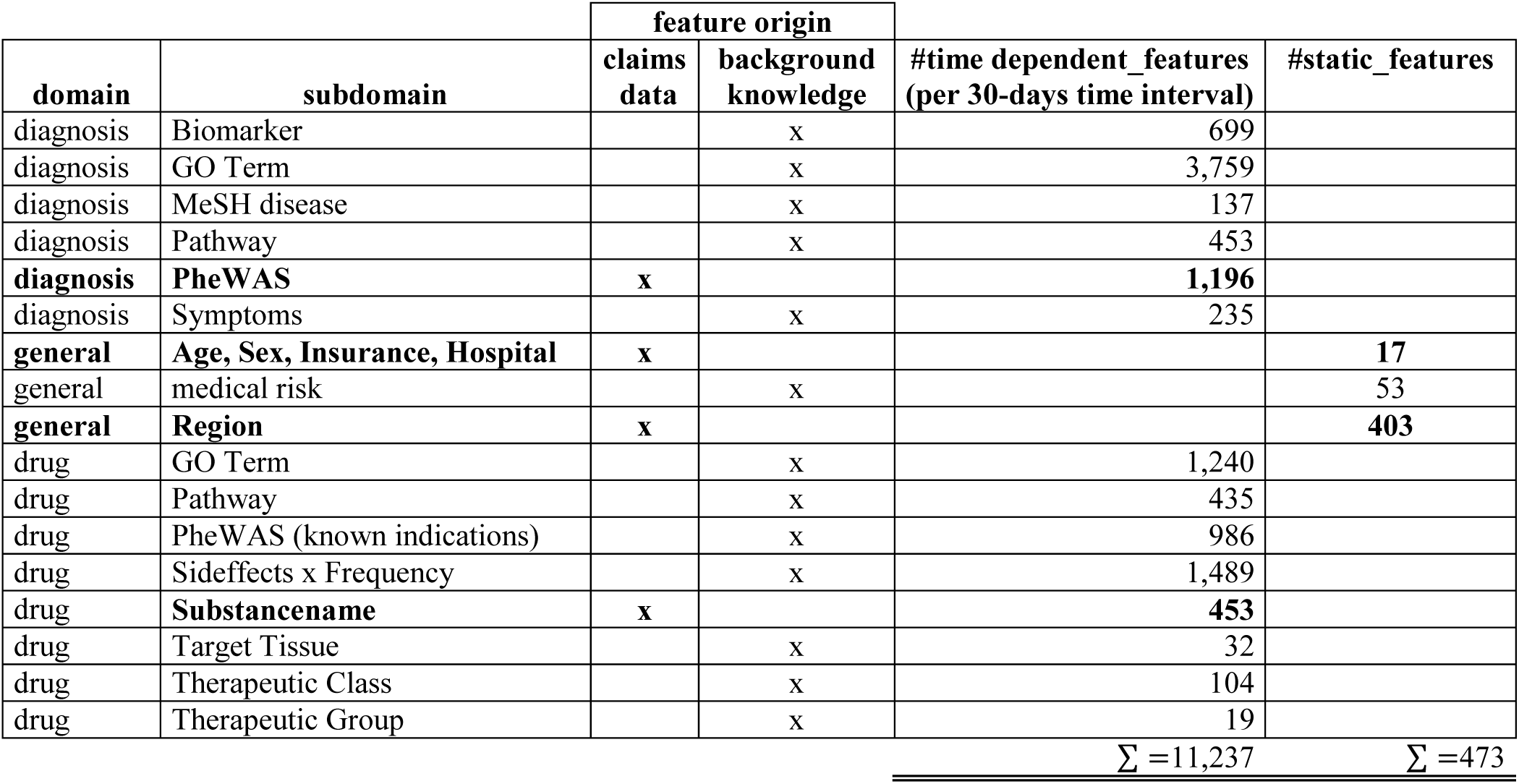
Number of features by domain, subdomain and feature origin (claims data or biomedical background knowledge).

| domain | subdomain | feature origin |  | #time dependent_features<br>(per 30-days time interval) | #static_features |
| --- | --- | --- | --- | --- | --- |
|  |  | claims data | background knowledge |  |  |
| diagnosis | Biomarker |  | x | 699 |  |
| diagnosis | GO Term |  | x | 3,759 |  |
| diagnosis | MeSH disease |  | x | 137 |  |
| diagnosis | Pathway |  | x | 453 |  |
| **diagnosis** | **PheWAS** | **x** |  | **1,196** |  |
| diagnosis | Symptoms |  | x | 235 |  |
| **general** | **Age, Sex, Insurance, Hospital** | **x** |  |  | **17** |
| general | medical risk |  | x |  | 53 |
| **general** | **Region** | **x** |  |  | **403** |
| drug | GO Term |  | x | 1,240 |  |
| drug | Pathway |  | x | 435 |  |
| drug | PheWAS (known indications) |  | x | 986 |  |
| drug | Sideeffects x Frequency |  | x | 1,489 |  |
| drug | **Substancename** | **x** |  | **453** |  |
| drug | Target Tissue |  | x | 32 |  |
| drug | Therapeutic Class |  | x | 104 |  |
| drug | Therapeutic Group |  | x | 19 |  |
| | | | | $\Sigma = 11,237$ | $\Sigma = 473$ |

**Figure 1:**
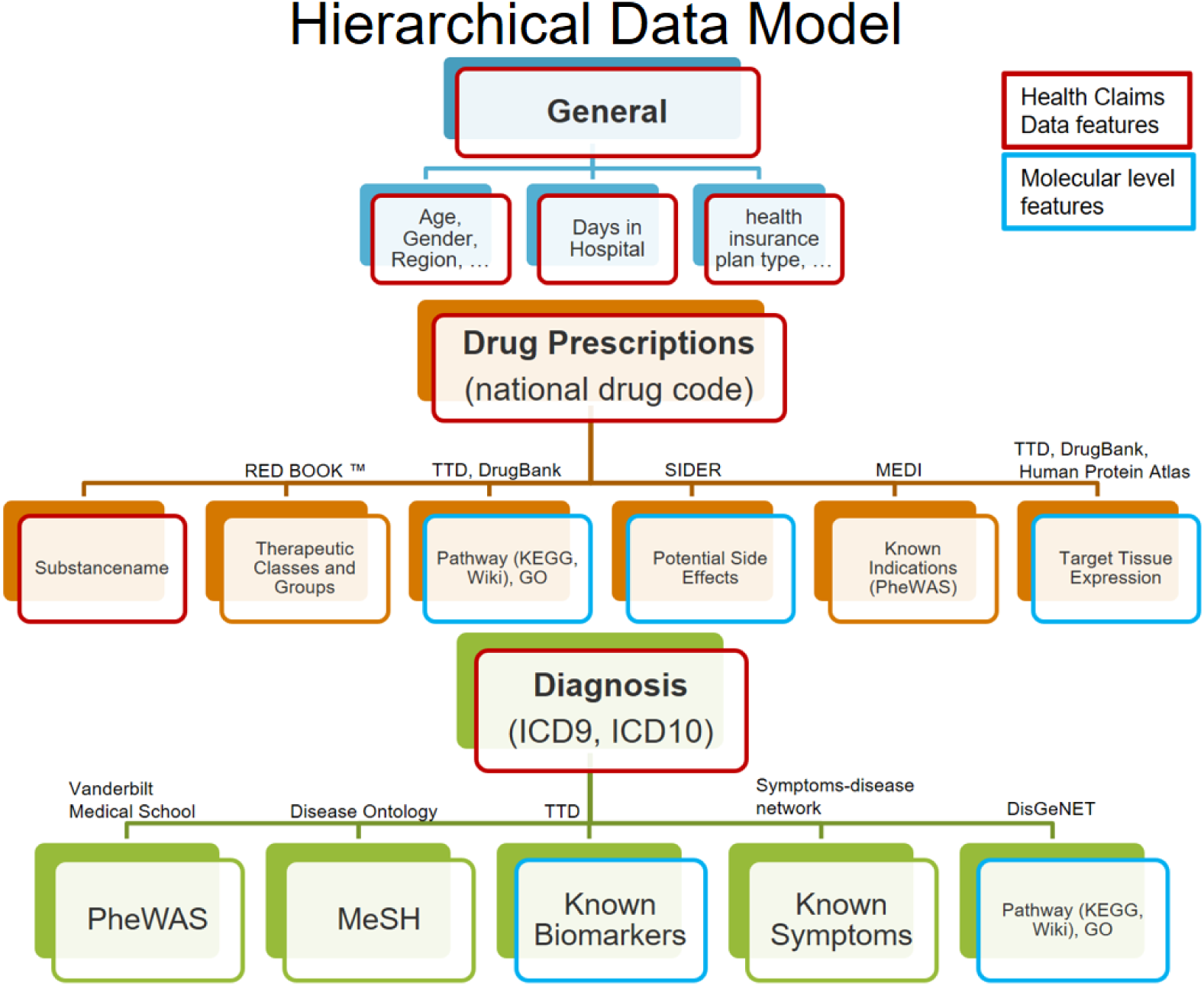
The hierarchical data model structures the data in 3 input domains (General, Drug, Diagnosis) and according subdomains. Features retrieved from the Claims Data are highlighted red, all others are augmented features (including those highlighted blue).

In this work we propose a multi-modal neural network architecture to reflect the specific structure of the augmented claims data, see **Figure 2**: In this architecture, each of the feature domains and sub-domains are initially treated as separate data modalities. Note that each feature derived from diagnosis and substance codes has an additional time stamp (30 days interval), i.e. each subdomain is a three-dimensional data cube. Each of these tensors is projected down to a lower dimensional representation via a bottleneck feed-forward architecture with 1 – 4 hidden layers, where the exact number of hidden layers and units per layer are treated as tunable hyperparameters in our framework, details in Supplements. In conclusion, at the first layers for each sub-domain, specific latent features are extracted in a non-linear manner from the original data and subsequently concatenated.

**Figure 2:**
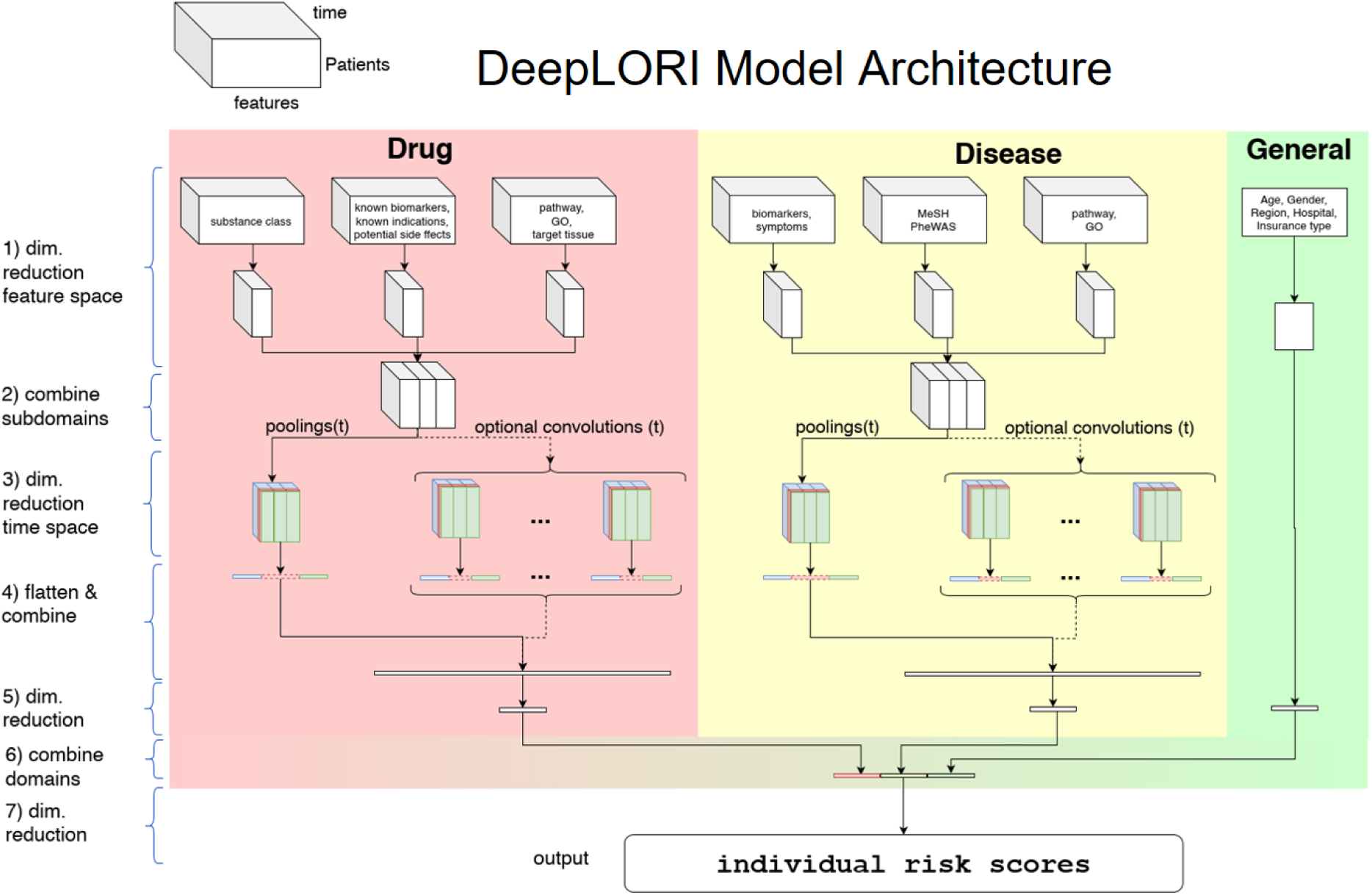
DeepLORI model architecture: The input is organized as a multi-modal data cube, which is sub-divided into several feature domains. Medication and diagnosis related features are time dependent, whereas features in the "General” domain are not. The three different colours at stage (3) symbolize the 3 different pooling/convolution kernel sizes.

After latent feature extraction, DeepLORI models the time dependency in the data within each feature domain. For that purpose we apply a pooling function (max or mean) over the entire time series, individually for each component of our one-hot-vector representations. The exact choice of the pooling function is a hyperparameter. In addition to pooling we allow for the application of different time convolutional kernels (with multiple filters per kernel), similar to a sliding window. Whether or not convolutional filters are applied, and which sizes these filters have, is again determined within hyperparameter optimization.

After modeling time dependency, there is another feedforward bottleneck structure (same tunable design as for the initial latent feature extraction) in our network. Finally, DeepLORI concatenates latent features extracted from each feature domain, and feeds them through the last feedforward bottleneck structure into one output unit, representing a patient specific comorbidity risk score. That means we have one DeepLORI model per comorbidity.

A more detailed view on the DeepLORI architecture, including an overview of all tunable hyperparameters can be found in the Supplementary material.

### 4.1.2 Loss Function

Let the training data denote as *D* = {(*x*_*it*_, *y*_*i*_, *δ*_*i*_) | *x*_*it*_ ∈ ℝ^*d*×*T*^, *y*_*i*_ ∈ ℝ, *δ*_*i*_ ∈ {0,1}, *i* = 1,2, …, *n, t* = 1,2, …, *T*}, where *T* is the maximum number of time stamps in the data (in our case 15 months) and *y*_*i*_ is the observed time of the first diagnosis of a comorbidity (in case the data is not censored) after index date or the maximum observed event free time. Moreover *δ*_*i*_ ∈ {0,1} is a binary variable indicating whether or not *y*_*i*_ is right censored (*δ*_*i*_ = 0) or not (*δ*_*i*_ = 1). Following (Katzman *et al*., 2018) we here use the negative partial log-likelihood of a Cox proportional hazard’s model (Cox, 1972) as a loss function for training DeepLORI:

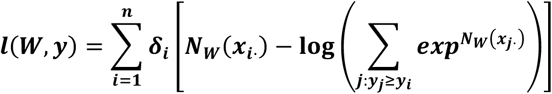

Here *N*_*W*_(·) denotes the risk score learned by DeepLORI, parameterized by weights *W*, and ***x***_***i***_. the medical history of patient *i*.

To avoid overfitting we regularize DeepLORI in multiple ways:

- We use drop-out units in the input and hidden layers.
- We perform batch normalization (Ioffe and Szegedy, 2015) before each activation function.
- We impose groupwise elastic net penalties for weights (Zou and Hastie, 2005).

The elastic net is an extension of the classical lasso algorithm (Tibshirani, 1996), which has originally been introduced in the context of generalized linear models. It combines an *ℓ*_1_ penalty of coefficients (like in lasso regression) with an *ℓ*_2_ penalty like in ridge regression (Hoerl and Kennard, 1970). The elastic net enforces a sparse regression model by jointly pushing coefficients towards zero via the *ℓ*_1_ penalty, i.e. there is a feature selection. At the same time the *ℓ*_2_ penalty promotes a joint selection of correlated features (Zou and Hastie, 2005). The idea of the elastic net can also be extended to neural networks. More specifically, by adding groupwise elastic net penalties we modify our training objective as:

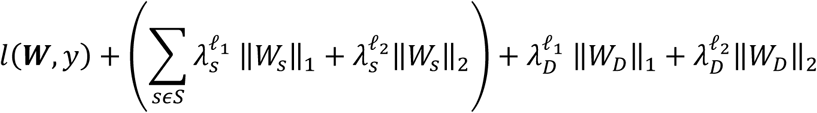

where *S* the set of feature subdomains. Furthermore, 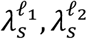 denote tunable hyperparamters, *W*_*s*_ refers to the set of weights connecting the input to the first hidden layer within feature domain *s*, and *W*_*D*_ are the weights of the connections feeding into the output layer.

### 4.1.3 Hyperparameters Optimization

A comprehensive overview of hyperparameters of DeepLORI can be found in the Supplements (Table S2). We performed Bayesian Hyperparameter Optimization (Bergstra, Yamins and Cox, 2013) to tune DeepLORI on the training data. Each candidate hyperparameter set was evaluated via a 5-fold cross-validation. Hyperparameter optimization was run for 100 trials per fold, a maximum number of 100 epochs per trial, or, if the cross-validated prediction performance did not increase within 10 sequential epochs. Prediction performance was measured via Harrel’s C-index (Harrell *et al*., 1982), which is a generalization of the area under receiver operating characteristic curve (AUC), frequently used for classification models.

### 4.1.4 Shapley Additive Explanations (SHAP)

One of the main criticisms of neural network based approaches is the difficulty to interpret them. Recently, (Lundberg and Lee, 2017) proposed a model agnostic game theoretic framework to address this issue. Briefly, the idea behind Shapley Additive exPlanations (SHAP) is that the relevance *Φ*_*i*_(*x*) of feature *i* on model output *f*(*x*) can be regarded as the average weighted difference between outputs from all possible models trained on (1) all subsets *S* of features including feature *i*, against (2) all subsets of features excluding feature *i*:

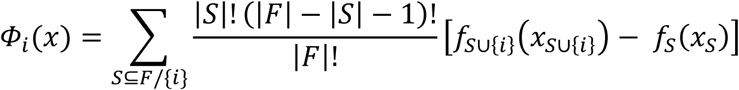

with *F* as the set of all features. The authors propose several local approximation techniques, which can circumvent the exact combinatorial calculation of *Φ*_*i*_(*x*), one which is specifically tailored towards neural networks (Deep SHAP). Deep SHAP effectively combines SHAP values calculated for smaller components of a neural network into SHAP values for the entire network. We refer to (Lundberg and Lee, 2017) for details. In this work we used SHAP to understand the impact of individual feature domains, sub-domains, and AEDs on the comorbidity risk score that we learned via DeepLORI. SHAP results in a patient specific score that may be positive (feature *i* increases *f*(*x*)) or negative (feature *i* decreases *f*(*x*)) compared to the average patient. In agreement to Lundberg et al. we considered the mean absolute values of the SHAP values per feature to score the overall impact of a variable. Moreover, we investigated the overall mean absolute SHAP values per feature domain and sub-domain, respectively. This is possible, because SHAP values are additive. That means the sum of two SHAP values can indeed be interpreted as the overall impact of the corresponding features.

In practice we found Deep SHAP too computationally costly when using our entire original dataset. We thus repeatedly subsampled 5% of our data with replacement (30 times) and re-calculated SHAP values. We checked the robustness of the approach via the variance of SHAP values.

### 4.2 Competing methods

We compared DeepLORI against several competing approaches:

1. Random Survival Forests (Ishwaran *et al*., 2008): In this earlier published approach (Gerlach, Lu and Fröhlich, 2017) we first combined claims data with biomedical knowledge (akin to this paper) and then used a window of fixed length (3 months) to summarize features via a max-pooling. Features encoding prescriptions and diagnoses within such a time window were concatenated, resulting into an overall number of around 165,000 features per patient. Subsequently we used maximum relevance minimum redundancy (mRMR) (Ding and Peng, 2005) to further reduce the number of features to 500 prior to Random Survival Forest (RSF) model training. For RSF model training we relied on R-package “ranger” (Wright and Ziegler, 2017). The number of decision trees was set to 5000, and the log-rank statistic was used as a split rule for nodes.
2. Stacked denoising autoencoders (SDA) followed by training a RSF (Miotto *et al*., 2016): In this approach initially an SDA was trained to extract features from the medical history of each patient (diagnosis and prescription codes as well as demographic information) in an unsupervised manner. The same SDA architecture as described in Miotto et al. was employed. After feature extraction for each of the 6 comorbidities a RSF was trained.
3. Kaplan-Meier (KM) estimator as “null model”. This approach does not use features of any individual. It only estimates the overall risk curve for a given comorbidity from the data and applies the same estimate to each patient. The purpose of this “null model” was to understand the added value of complex machine learning models.

### 4.3 Evaluation Approach

DeepLORI was compared with competing methods within a 5-fold cross-validation scheme using the exactly same data splits of the original dataset. Hyperparameters were only tuned on the respective training data, resulting in a nested cross-validation scheme for DeepLORI. We used Uno’s C-index (Uno *et al*., 2011) as a performance measure:

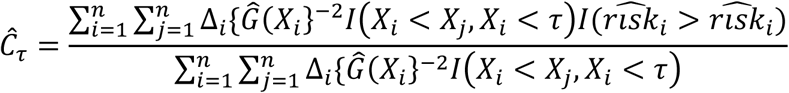

Uno’s C-index is a consistent estimator of the concordance index for a population that is independent of censoring. It satisfies this requirement for censored populations using two “tricks”, first by applying a “inverse probability weighting” schema using the censoring distribution estimated with *Ĝ*(·) (e.g. the Kaplan-Meier estimator), second by evading instable tail parts for times ≥ *τ* of the estimated survival function with a prespecified time point *τ* as constraint. We refer to (Uno *et al*., 2011) for further details.

In addition, we evaluated DeepLORI on our external validation data. This validation was done separately in two different ways:

1. Follow-up of existing patients: We selected patients that had already been in our original dataset, but for which a right censoring was observed.
2. New patients: We evaluated DeepLORI on patients that had no records in the original dataset.

In both cases we recorded Uno’s C-index over the entire time series and as a function of time (named *AUC*(*t*) in the following) to measure the prediction performance.

## 5 Results

### 5.1 DeepLORI Outperforms Competing Methods

Despite high censoring rates all 6 comorbidities could be predicted rather accurately by DeepLORI, and the 5-fold cross-validated performance of Uno’s C-index ranged from 71% for overweight and obesity up to 77% for stroke & ischemic attacks (**Figure 3, Table S3**). At the same time the Kaplan-Meier estimator (i.e. the “null”-model without any features) was consistently at chance level (50% Uno’s C-index), indicating that all of our tested machine learning models (DeepPatient, DeepLORI and MRMR + RSF) extracted relevant predictive signal from the data. At the same time DeepLORI showed significantly higher C-indices than all competing methods.

**Figure 3:**
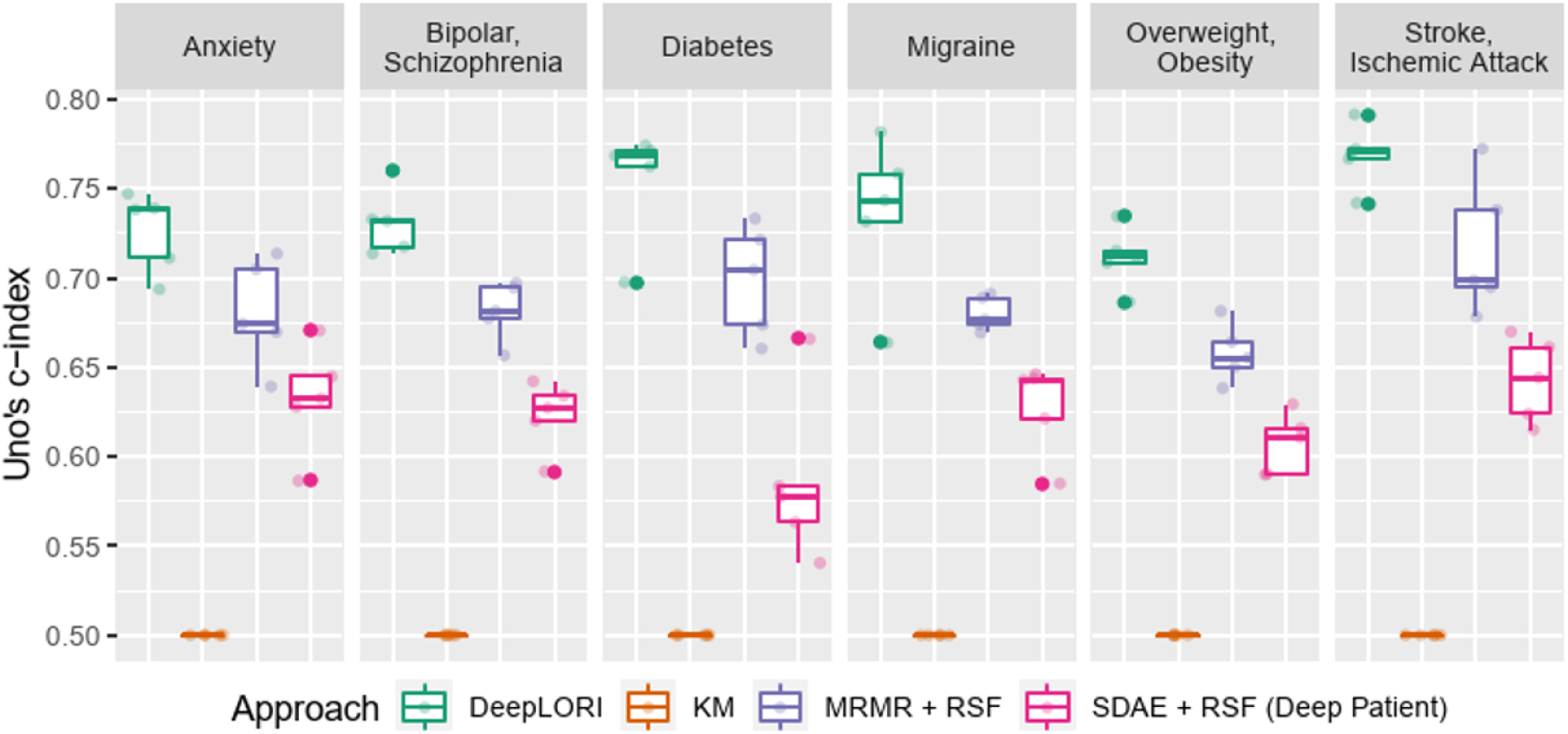
5-fold (nested) cross-validation test set performance benchmark of **Deep LORI** (green) vs. competing methods; **MRMR**: Minimum Redundance Maximum Relevance feature selection; **SDAE**: Stacked Denoising Autoencoder; **RSF**: Random Survival Forest; **KM**: Kaplan-Meier estimator.

### 5.2 DeepLORI Shows Stable Prediction Performance on External Validation Data

Evaluation of DeepLORI on the external validation data showed roughly comparable C-indices to those observed on the original data when focusing on the follow-up of the ∼15,000 patients, which had medical history in the original data (**Figure 4**). This highlights that DeepLORI, despite of high censoring rates in the original data, was not overfitted. C-indices for new / so far unseen patients (n = ∼97,000) in the external validation data were around 6% lower (**Table S3**).

**Figure 4:**
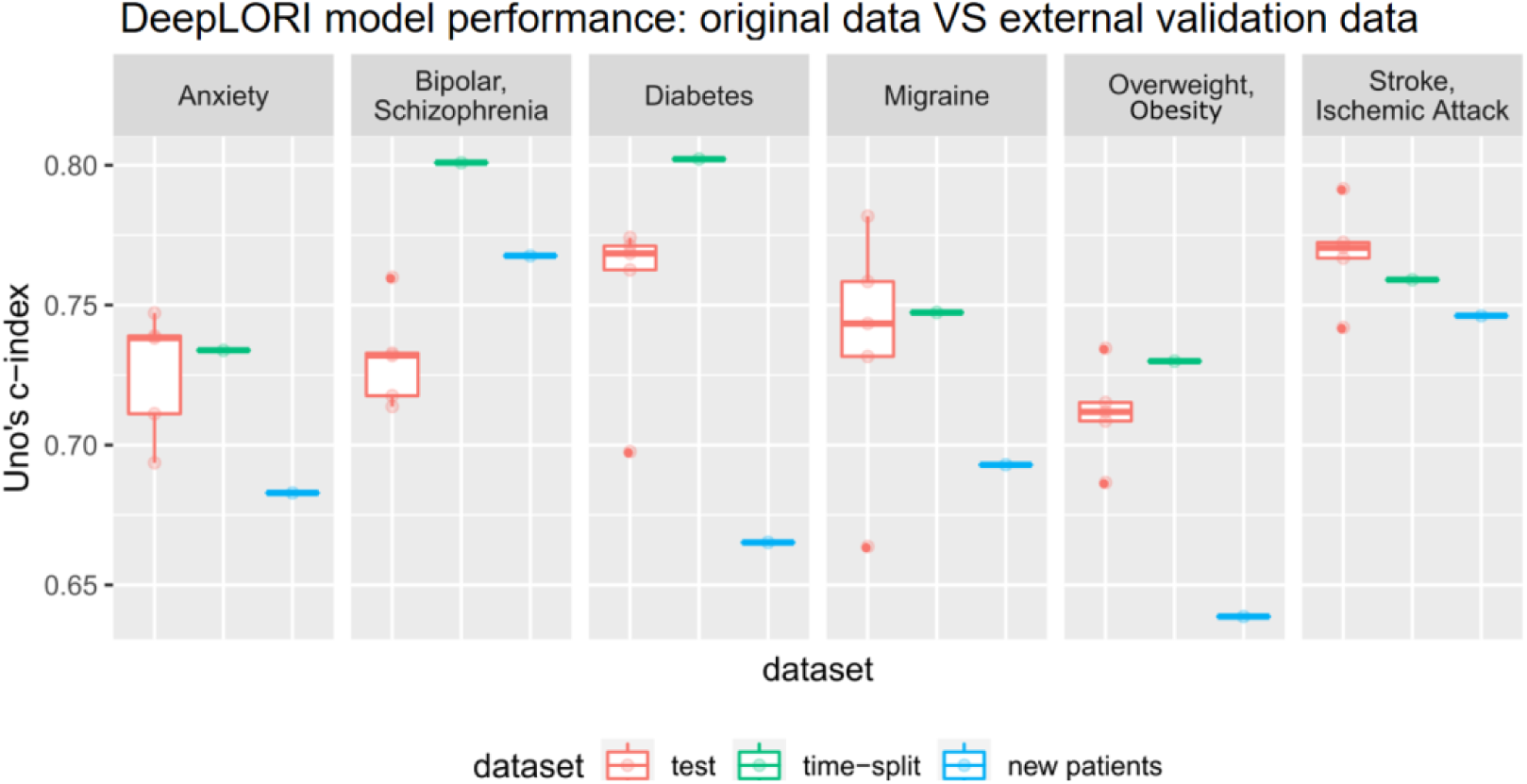
Prediction performance of Deep LORI on external validaton data. Green: time-split validation, i.e. follow up of patients included into training data. Blue: Prediction performance on new patient data. Red: 5-fold cross-validated prediction performance on original data for comparison purposes.

When investigating the *AUC*(*t*) we found a rather stable prediction performance for all comorbidities over time (**Figure 5**). Remarkably this held true for a time interval of up to 6 years after initial diagnosis of epilepsy, and it was true for the follow-up of existing patients as well as for new patients, again highlighting the fact that DeepLORI generalizes rather well.

**Figure 5:**
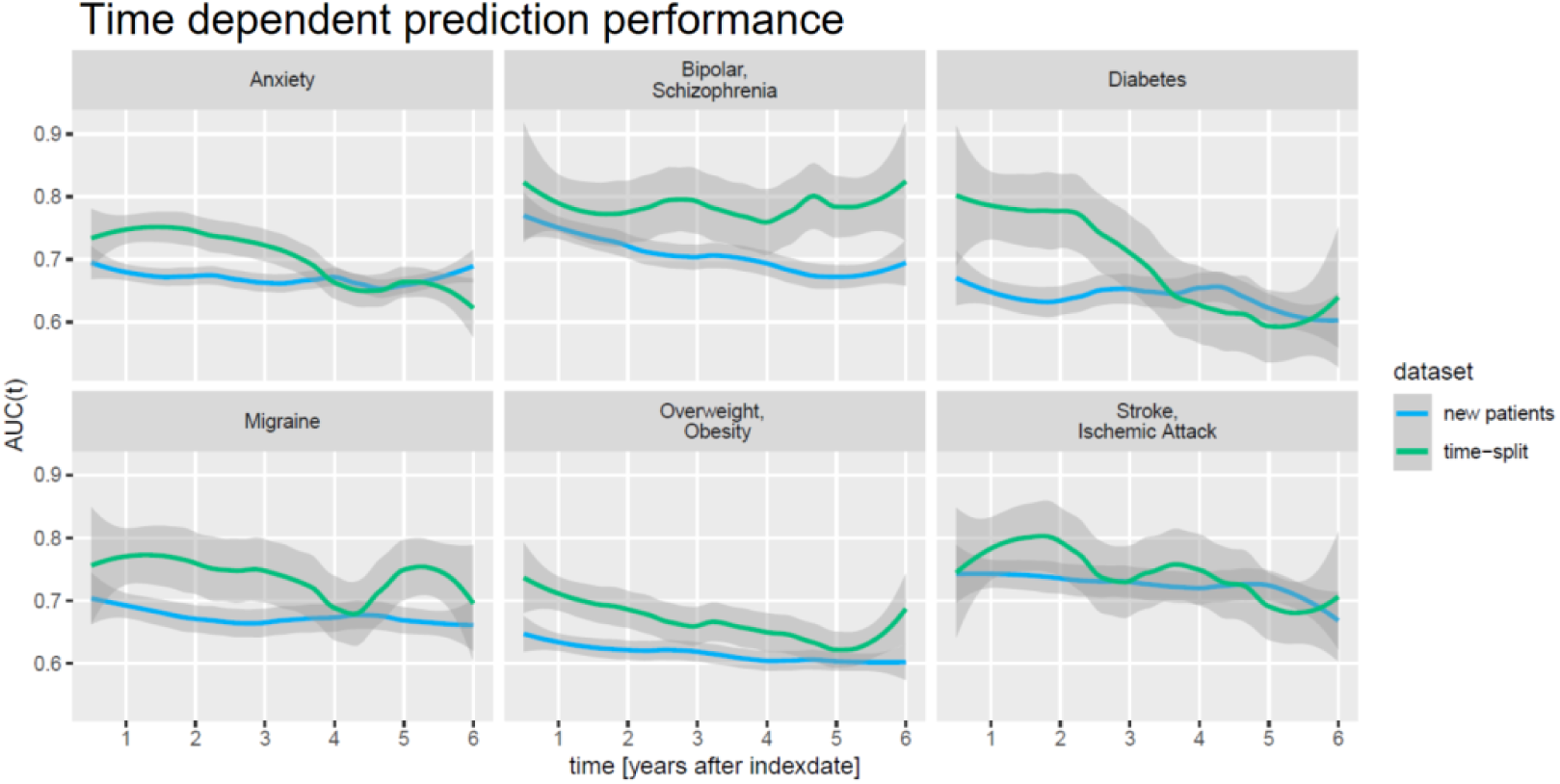
Time-dependent prediction performance (AUC(t)) of DeepLORI on external validation data.

### 5.3 DeepLORI Models are Explainable

We next investigated the relevance of feature domains via SHAP in DeepLORI models trained on the entire original dataset. This analysis highlighted for most comorbidities the relevance of features derived via augmentation of the original data with additional information (**Figure 6**): For example, in the model predicting anxiety augmented features made up around 55% of the total feature importance, for migraine we found 43%. Notably, among augmented features the “medical risk” subdomain, covering various unspecific comorbidity indices derived from ICD diagnosis codes (Charlson *et al*., 1987; Romano, Roos and Jollis, 1993; Lee *et al*., 1999; Schneeweiss *et al*., 2003; Boersma *et al*., 2005; Quan *et al*., 2005; Sessler *et al*., 2010; Sigakis, Bittner and Wanderer, 2013) using the R-package “medicalrisk” (McCormick and Joseph, 2020) in the medical history of epilepsy patients, had a significant impact (**Figure 7**), suggesting that the risk of developing any of our six comorbidities increases with a generally worse medical condition upfront. Importantly, none of the comorbidity indices are specific to any of the six comorbidities focused by DeepLORI.

**Figure 6:**
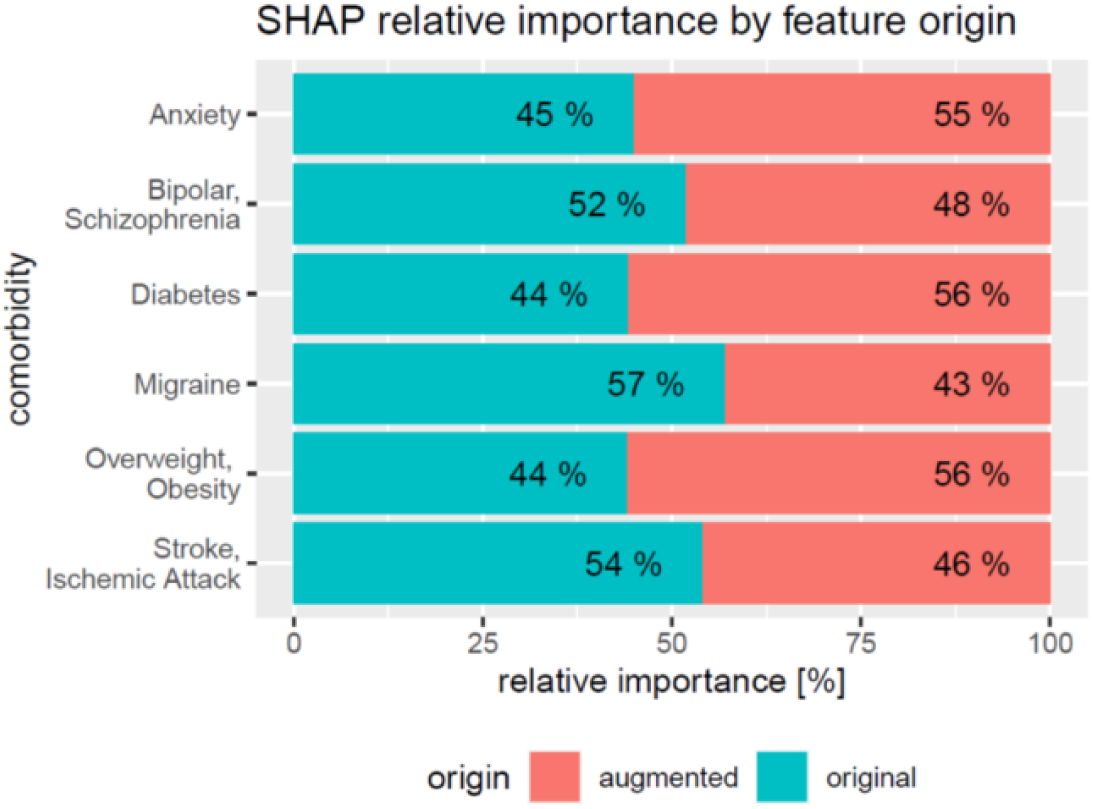
Relative impact of original vs augmented features in DeepLORI models for 6 different comorbidities

**Figure 7:**
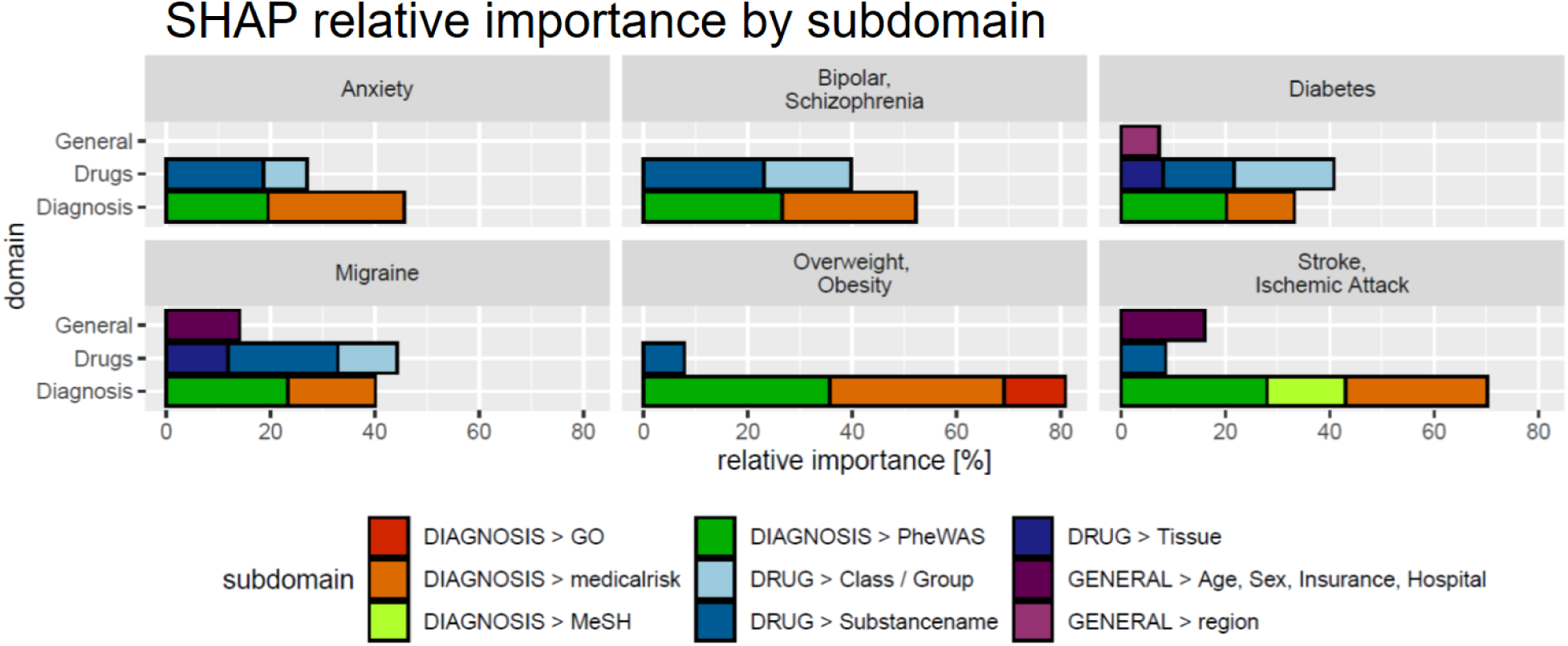
SHAP values by feature subdomain. Subdomains with a cumulative relative importance ≤ 5% are not shown.

In the following we discuss one of the six DeepLORI models in more detail, namely the one for migraine **(Figure 8**). Figures related to DeepLORI models for the other five comorbidities can be found in the Supplementary Material (**Figures S1 – S6**). According to SHAP analysis the most relevant features in the DeepLORI model for migraine relate to the prior existence of headaches and the use of drugs for the nervous system, which are typically used to treat headaches. In addition, many drugs used for treating headaches are known to affect the liver (Mathew and Klein, 2019; Valade, 2019). It is known that females are more affected by migraine than males, and that migraine is age dependent (Victor *et al*., 2010). The anti-epileptic drug (AED) topiramate, known to be well tolerated by this group of patients (Spritzer, Bravo and Drazkowski, 2016; Silberstein, 2017), ranks among the top 15 most relevant features: **Figure 9** shows the marginal dependency of DeepLORI model predictions on AED prescription frequency, suggesting that patients treated with topiramate are slightly more likely to be diagnosed with migraine later on compared to those without such treatment in the past. In fact, topiramate is often used as a preventive treatment for migraine (Spritzer, Bravo and Drazkowski, 2016), suggesting that patients treated with this AED are often considered at risk of developing migraine by their treating physician. Indeed, many of these patients eventually receive this diagnosis.

**Figure 8:**
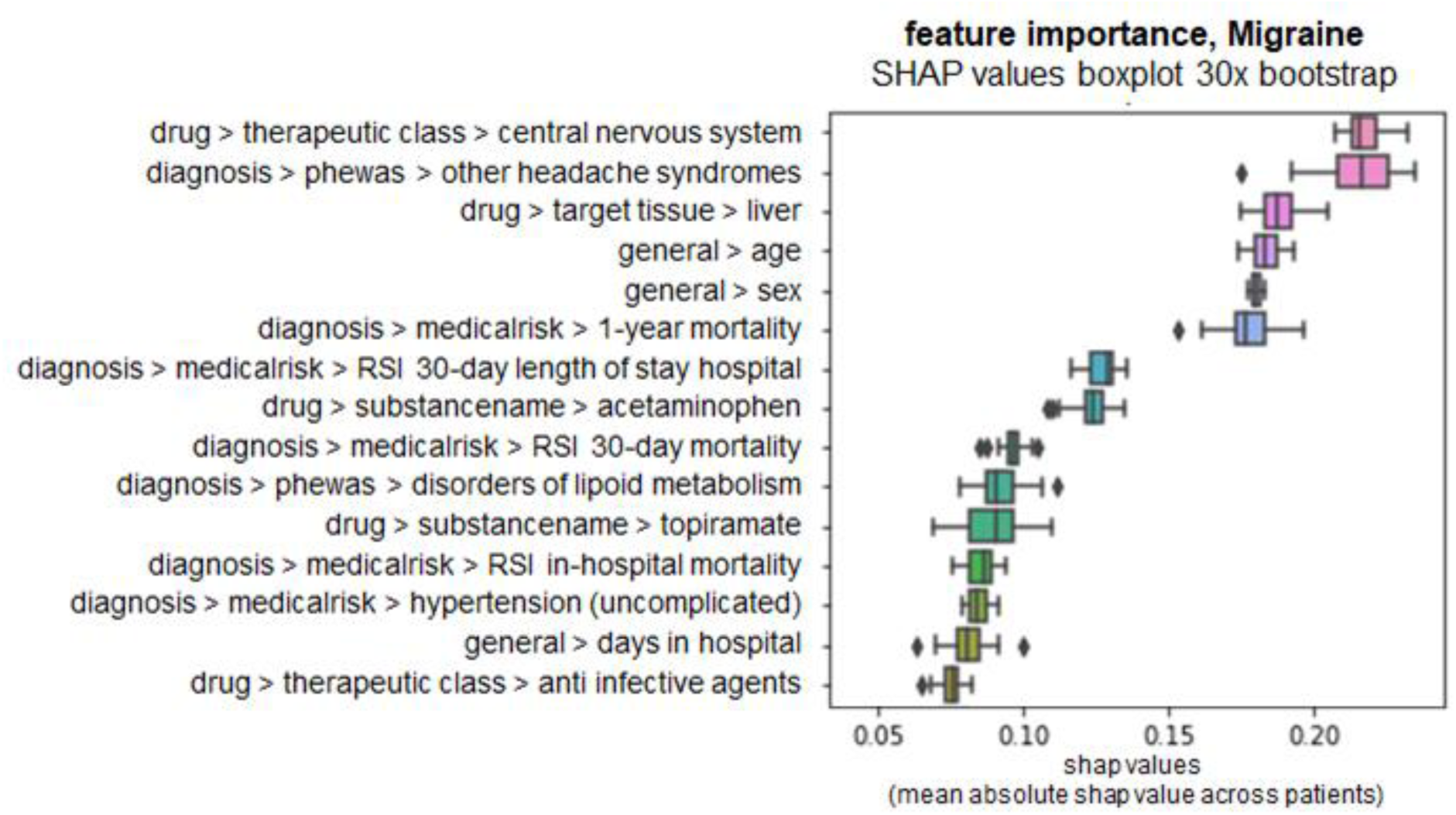
SHAP values of top 15 most relevant features in the DeepLORI model for migraine, rank by mean absolute values. x-axis: SHAP values, higher values = higher importance. y-axis: features ranked by importance, annotated with their domain and subdomain membership (domain > subdomain > feature),

**Figure 9:**
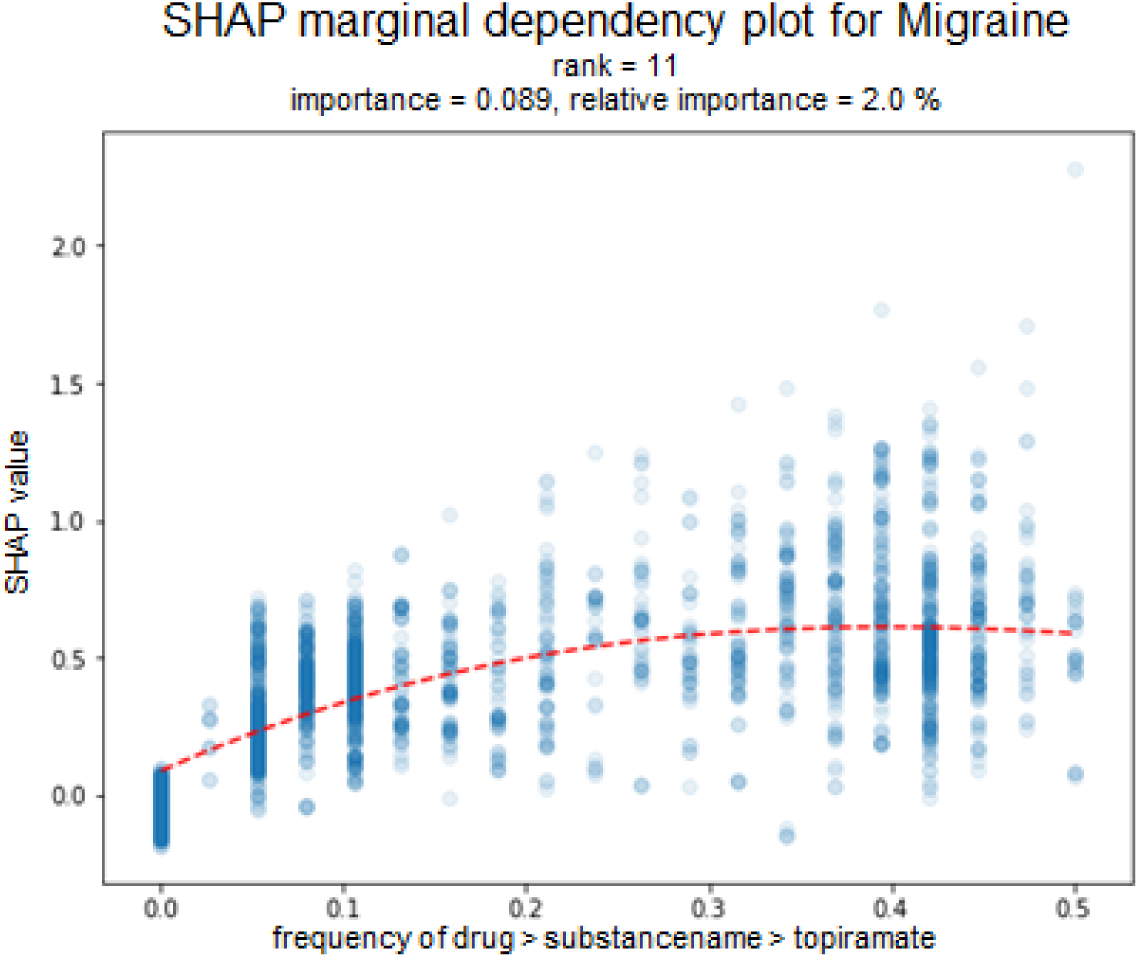
The plot shows the marginal dependency of the frequency of topiramate prescription on the predicted risk on later migraine diagnosis. One dot represents one patient; x-axis: prescription frequency (0% = never observed, 100% = observed in every month of a patient’s medical history); y-axis: change in SHAP-value (=change in individual hazard rate). Marginal dependency plots for each comorbidity for the top-5 features can be found in the Supplemental Material (Figures S7-S12).

Another interesting finding from the SHAP analysis of our model is the influence of disorders of the lipoid metabolism on migraine risk. Associations between lipid levels and migraine have been reported in (Rist, Tzourio and Kurth, 2011) and (Onderwater *et al*., 2019). Moreover, the metabolic syndrome and migraine have been associated to each other (Sachdev and Marmura, 2012). It is important to highlight at this point that SHAP analysis does not provide a causal explanation, though.

To further exemplify the possibility of interpreting our DeepLORI models on the level of individual patients we depict in **Figure 10**. SHAP values for two randomly selected patients with high and low risk for developing stroke or ischemic attacks, respectively. As expected, the low risk patient is young and has no diagnosis of hypertension or disorder of the metabolic system. In contrast, the high risk patient is an older person with hypertension who lives in Texas. In fact, significant regional differences in the risk for strokes have been reported throughout the US (Howard *et al*., 2007), and Eastern Texas belongs to the so-called "Stroke Belt” (Karp David N. *et al*., 2016).

**Figure 10:**
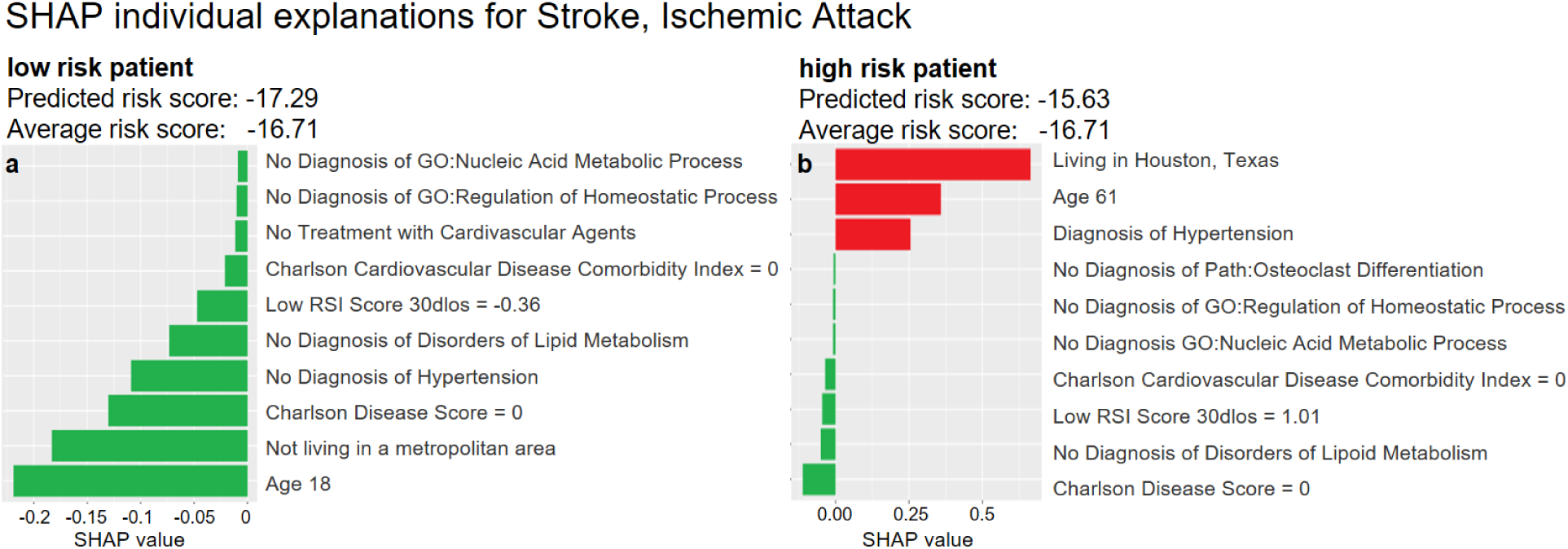
Examples of SHAP explanations for a patient predicted at low risk for stroke & ischemic attacks (left) and a high risk patient (right). Red (green) bars indicate higher (lower) risk compared to the average patient: The hazard ratio^3^ of the low risk patient (left) is around 56% of that of an average patient 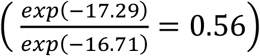. The hazard ratio of the high risk patient (right) is around 2.94 times that of an average patient 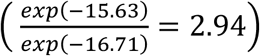.

## 6 Discussion and Conclusions

Precision medicine has the vision to bring the right treatment to the right patients. Precision medicine is strongly dependent on machine learning. At present precision medicine is only an emerging reality. Several reasons can be identified (Fröhlich *et al*., 2018; Miotto *et al*., 2018; Xiao, Choi and Sun, 2018; Kwak and Hui, 2020): 1) lack of the right data in sufficient quantity; 2) insufficient validation; and 3) difficulties in interpreting complex ML models, which is by itself a prerequisite for generating the necessary confidence in using such models. Realization of precision medicine will only be possible, if all these aspects are addressed jointly. In this context it is essential that ML models can be used in a cost effective and practical manner. Hence, clinical routine data are of extreme relevance and are gaining more and more attention (Weiss *et al*., 2012; Peissig *et al*., 2014; Miotto *et al*., 2016; Choi *et al*., 2016; Rajkomar *et al*., 2018; Harutyunyan *et al*., 2019). Administrative claims data constitute an important source of such clinical routine data. They principally exist in large quantities and allow for obtaining insights into the longitudinal medical history of individual patients under real world conditions. However, these data have not been collected for research purposes. First of all, coding of diagnoses into ICD codes is not unique and mostly done for maximizing economic reasons rather than for providing a precise medical description. Different ICD codes can be used for similar diagnoses, and the relationship between different medical conditions is consequently not always uniquely resolvable from their distances in the ICD ontology. Second, it should be noted that ICD only reflects the medical symptom level, which should not be confused with the biological relationship between disorders. Third, the time of diagnosis encoding might not correspond to the actual appearance of the medical condition. Fourth, it is unclear whether patients take the prescribed medication. Finally, the nature of irregular time series data, different for each patient, imposes specific challenges for data analysis.

In this work we tried to address these challenges by a) mapping ICD codes to PheWAS codes that are at higher granularity; b) augmenting the original data with further information from biological databases; and c) proposing a specific multi-modal neural network architecture (DeepLORI). We demonstrated that DeepLORI can predict six common comorbidities of epilepsy patients with higher C-index than several competing methods. We performed a rigorous cross-validation plus an external validation to assess our model, demonstrating that DeepLORI allows for reliable predictions of comorbidity risks up to six years in advance. We showed that with the help of SHAP and our data augmentation approach it is possible to make DeepLORI based predictions explainable, even on the level of individual patients. From our perspective this is of great importance for generating confidence in ML based solutions in medicine.

From a medical perspective we see the value of our work in the potential for much earlier identification of epilepsy patients at risk of developing different comorbidities. For example, a patient at high risk of developing diabetes type 2 should consider losing weight and regularly check insulin levels. A patient at high risk of developing psychiatric disorders might consider early consultation with a psychiatrist. Hence, risk models could be a way to eventually move towards preventive medicine.

Further applications of our work could lie in addressing the high subject-to-subject variability in epilepsy: Based on the comorbidity risk profile learned by DeepLORI one might be able to identify subgroups of patients with more homogenous disease progression, potentially opening up opportunities for developing more personalized therapies in the future.

## Data Availability

The data analyzed in this study is subject to the following licenses/restrictions: We used administrative health claims data provided by IBM Truven Health Analytics. Data access has to be requested from IBM. Requests to access these datasets should be directed to 
https://www.ibm.com/products/marketscan-research-databases, https://www.ibm.com/watson-health/about/truven-health-analytics .

## 7 Abbreviations

AED: Anti Epileptic Drug
SDA: Stacked Denoising Autoencoder
MRMR: Minimum Redundance Maximum Relevance
RSF: Random Survival Forest

## 8 Acknowledgements

We like to thank Linda Kalilani, Babak Boroorjerdi for helpful discussions and support of the entire project.

## 10 Author Contributions

Designed and supervised the project: HF; extracted the data: CL, HK, KH; analyzed the data: TL, JdJ; performed experiments: TL; interpreted the data and results: TL, HF; drafted the manuscript: TL, HF. All authors have read the manuscript and agreed to its content.

## Footnotes

1 Surname changed from Gerlach to Linden in 2018

2 http://www.tdrdata.com/ipd/ipd/ICD10ToICD9List

3 Hazard ratios computed similar to Cox proportional hazard’s model (Cox, 1972).

