## supplemental for "An Explainable Multi-Modal Neural Network Architecture for Predicting Epilepsy Comorbidities Based on Administrative Claims Data"

### 3 DATA

#### 3.2 DEFINITION OF FOCUSED COMORBIDITIES AND COMPILATION OF TRAINING DATA

| Comorbidity | PheWAS Code description |
|---|---|
| Anxiety | "Anxiety disorder",<br>"Generalized anxiety disorder",<br>"Anxiety, phobic and dissociative disorders",<br>"Agorophobia, social phobia, and panic disorder" |
| Bipolar, Schizophrenia | "Bipolar",<br>"Schizophrenia and other psychotic disorders" |
| Diabetes | "Type 2 diabetes" |
| Migraine | "Migraine",<br>"Migrain with aura" |
| Overweight, Obesity | "Overweight", "Obesity" |
| Stroke, IschemAttack | "Ischemic stroke",<br>"Transient cerebral ischemia" |

**Table S 1:** Overview of comorbidity defintions using PheWAS.

[1] Last name changed from Gerlach to Linden in 2018

##### 4.1.3 Integration of Biological Background Knowledge into Claims Data

We used DisGeNET (Piñero *et al.*, 2017) to retrieve for each reported diagnosis in our data disease associated genes. Enrichment of Gene Ontology (GO) biological processes ('The Gene Ontology Consortium', 2004), KEGG (Kanehisa, 2000) and Wiki pathways (Slenter *et al.*, 2018) was then estimated via a conditional hyper-geometric test using GOstats (Falcon and Gentleman, 2007) with a tail area based false discovery rate (Strimmer, 2008) cutoff of 5% for GO and 20% for pathways. Furthermore, known disease biomarkers and symptoms were obtained from the Therapeutic Target Database (TTD) (Yang *et al.*, 2016) and the human symptoms-disease network (Zhou *et al.*, 2014). Similarly, medications in the claims data were mapped to known targets via TTD and DrugBank (Wishart, 2006) via text matching of substance names, and information about tissue expression of drug targets was obtained from the Human Protein Atlas (Uhlen *et al.*, 2015). Potential side effects of drugs and their likelihoods were retrieved from SIDER (Kuhn *et al.*, 2016), again by application of text matching of substance names. We obtained information about likely indication areas of drugs from MEDI (Wei *et al.*, 2013). Notably, only indication areas that were marked as "high confidence" by the MEDI authors and were mentioned in at least 10 PubMed articles were used. Finally, medications were mapped to substance classes and groups via the RED BOOK™ database. Further details are described in (Gerlach, Lu and Fröhlich, 2017).

##### 4.1.4 Hyperparameters Optimization

| hyperparameter | sample space | lower bound | upper bound | choice | comment |
|---|---|---|---|---|---|
| architecture | | | | | |
| #layers | choice | | | {1, 2, 3, 4} | |
| #units/layer shrinkage | conditional on #layers | | | 1 layer: {1%, 0.1%}<br>2-4 layers: {10%, 1%} | |
| activation function | choice | | | tanh, selu | |
| initializer | condtitional on activation function | | | tanh: {glorot normal, orthogonal}<br>selu: {lecun normal} | |
| pooling method | choice | | | {max, mean} | 3 windows: [3,6,17] |
| convolution | choice | | | {yes, no} | 5 filters per window. 3 windows: [3,6,17] |
| regularization | | | | | |
| L1 | log_uniform | 1.00E-02 | 1.00E-07 | | |
| L2 | log_uniform | 1.00E-02 | 1.00E-07 | | |
| dropout input | uniform | 0% | 10% | | |
| dropout hidden | uniform | 0% | 50% | | |
| batch normalization | choice | | | {yes, no} | allow "no" for selu |

| | | | | | |
|---|---|---|---|---|---|
| batch size | choice | | | {64,128,256,512} | (Keskar *et al.*, 2016) |
| training configuration | | | | | |
| learning rate | log_uniform | 1.00E-01 | 1.00E-04 | | |
| optimizer | choice | | | {SGD nesterov, RMSProp, Nadam} | |

**Table S 2:** Overview of hyperparameters tuned with Bayesian Hyperparameter Optimization. Hyperparameters were sample from continuous distributions (uniform, log-uniform), discrete (#layers, #units/layer, batch size) and categorical (all others) spaces. Some subspaces are conditional on the sampling outcome of another hyperparameter, e. g. the initializer depends on the activation function and the width (#units/layer shrinkage) depends on the depth (#layers).

**Details on Modeling Time Dependency**

In our case max pooling corresponds to a mapping from $\{0,1\}^w \rightarrow \{0,1\}$ performing a qualitative aggregation and mean pooling from $\{0,1\}^w \rightarrow [0,1]$ to perform a quantitative aggregation across a patients medical history. Where $w$ are the number of timepoints per feature for a given window size, e.g. $w = 3$ means that data from 3 consectuive timpoints are aggregated.

#### 5.1 DEEPLORI OUTPERFORMS COMPETING METHODS

| | Uno's c-index | | | |
|---|---|---|---|---|
| | original data | external validation data (n=112,755) | | Difference |
| comorbidity | test set (n=19,510) | time-split (n=15,276) | new patients (n=97,479) | time-split VS new patients |
| Anxiety | 0.73 (0.02) | 0.73 | 0.68 | 0.05 |
| Bipolar, Schizophrenia | 0.73 (0.02) | 0.80 | 0.77 | 0.03 |
| Diabetes | 0.75 (0.03) | 0.80 | 0.67 | 0.13 |
| Migraine | 0.74 (0.04) | 0.75 | 0.69 | 0.06 |
| Overweight, Obesity | 0.71 (0.02) | 0.73 | 0.64 | 0.09 |
| Stroke, Ischemic Attack | 0.77 (0.02) | 0.76 | 0.75 | 0.01 |

**Table S3:** Prediction performances (Uno's c-index) of the observed endpoints. Original data: 5x5-fold nested cross-validation; mean = mean performance of the best model; *se = standard error of the mean. External validation data: Prediction performance based on the final models.

#### 5.2 Interpretation of DeepLORI Models via SHAP

##### BOXPLOT 30X BOOTSTRAP TOP-15 FEATURES

The boxplots in this section show the distribution of shap values: We repeatedly subsampled 5% of our data with replacement (30 times) and re-calculated SHAP values. We checked the robustness of the approach via the variance of SHAP values.

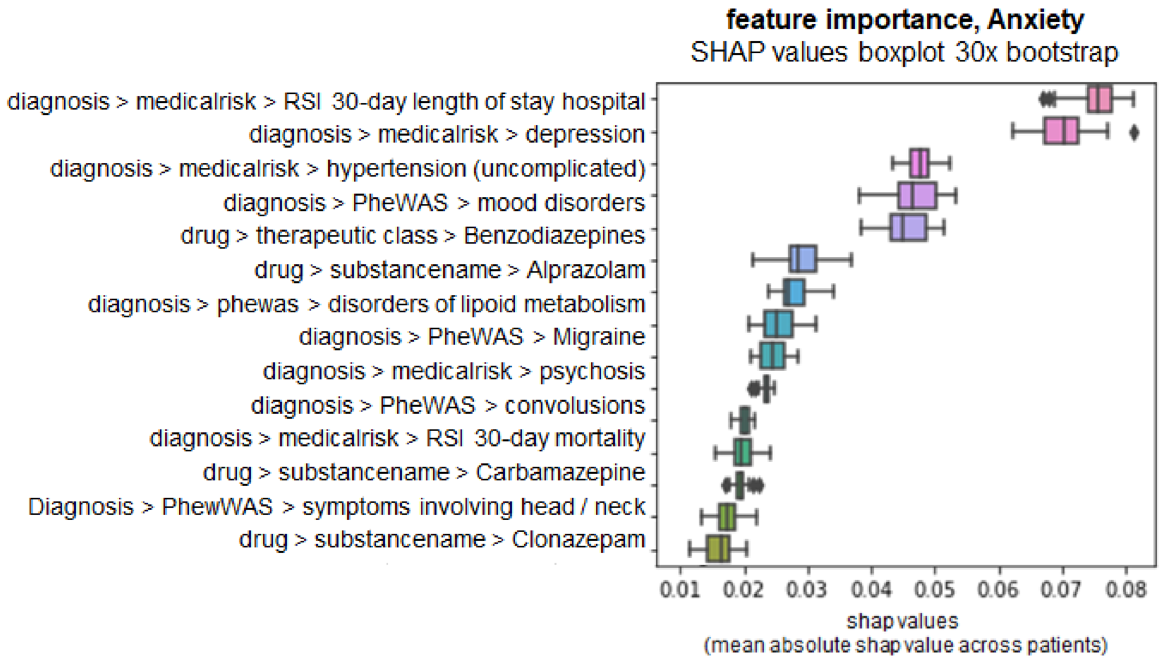


**Figure S 1:** SHAP boxplot feature importance Anxiety

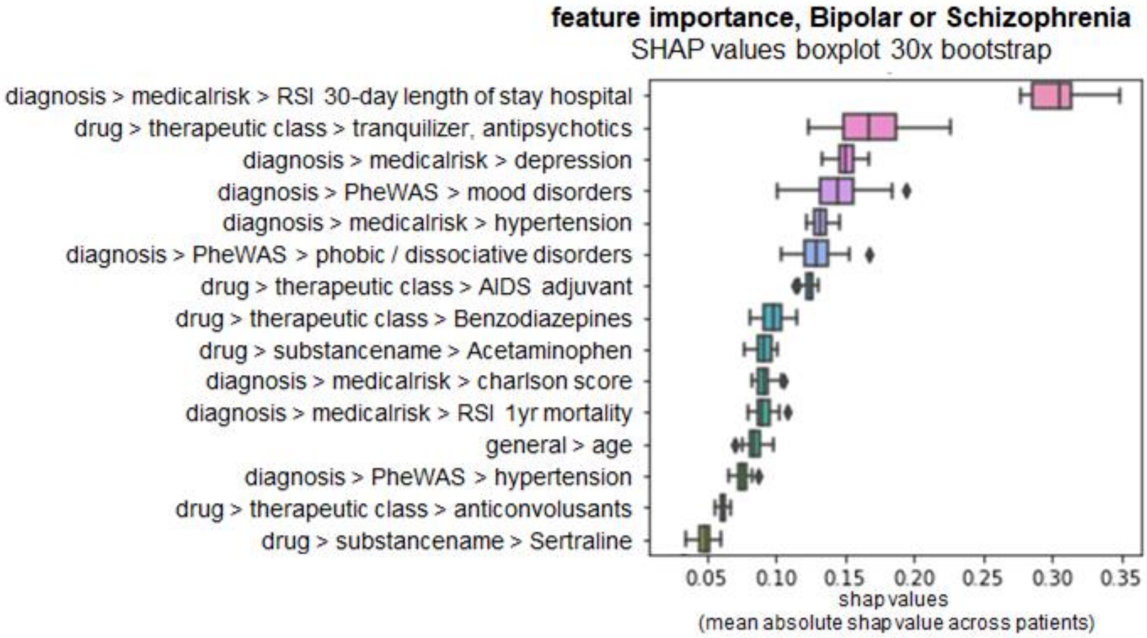


**Figure S 2:** SHAP boxplot feature importance Bipolar or Schizophrenia

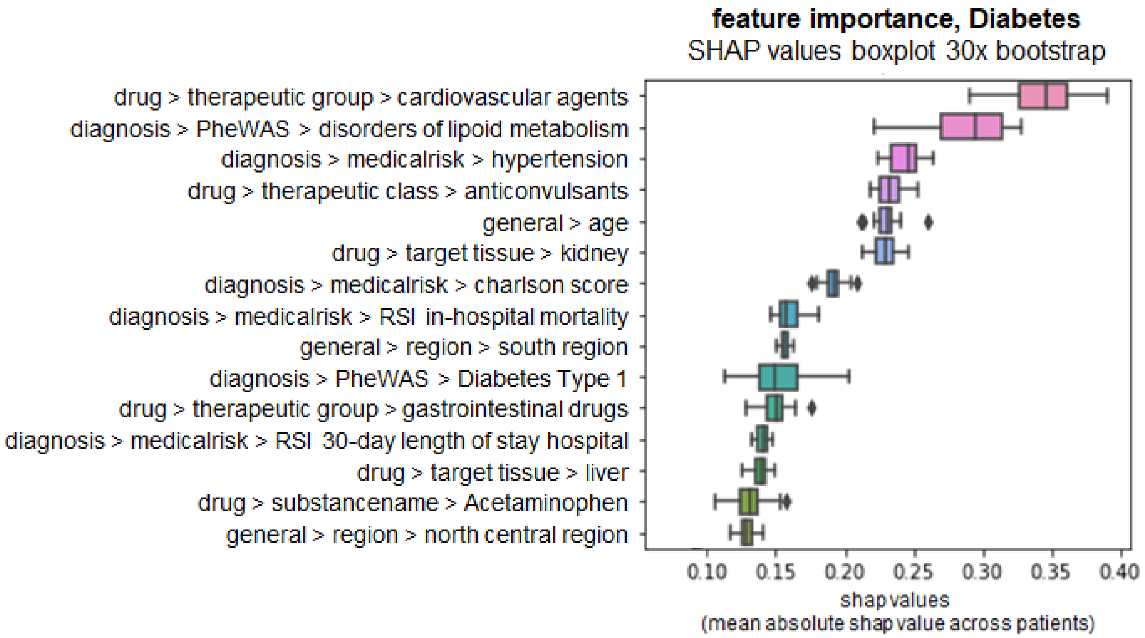


**Figure S 3:** SHAP boxplot feature importance Diabetes type 2

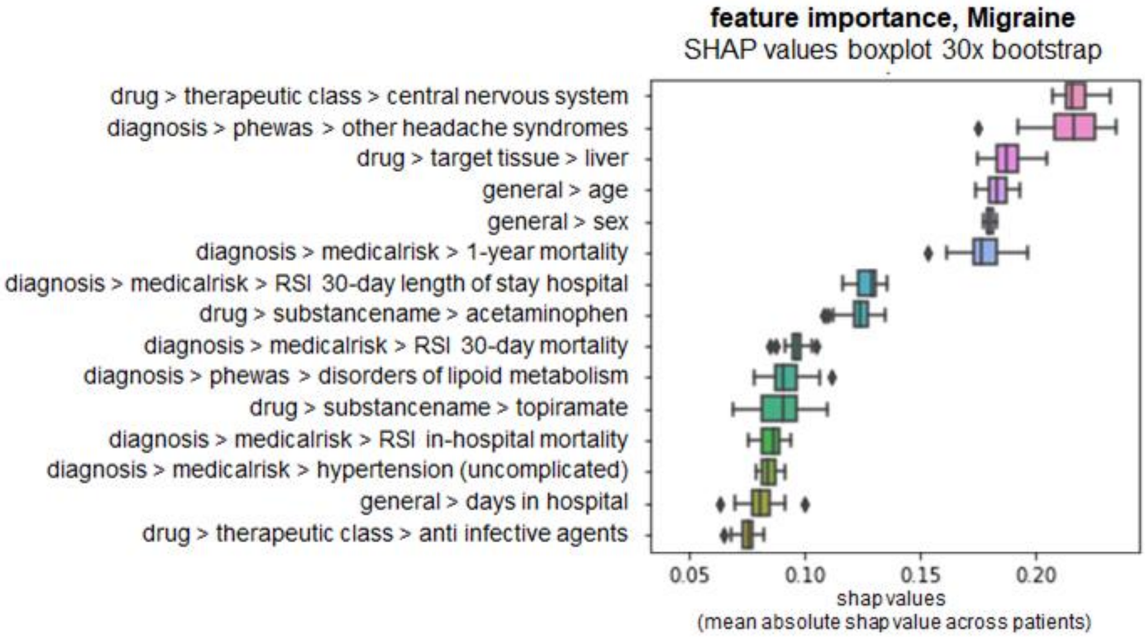


**figure S 4:** SHAP boxplot feature importance Migraine

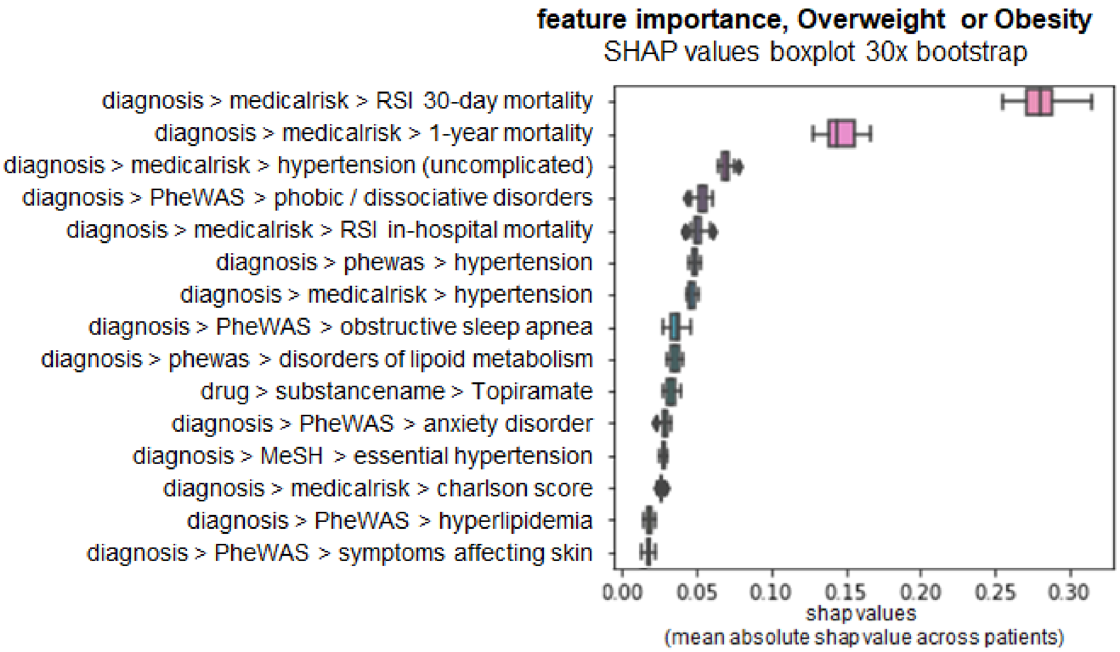


**figure S 5:** SHAP boxplot feature importance Overweight or Obesity

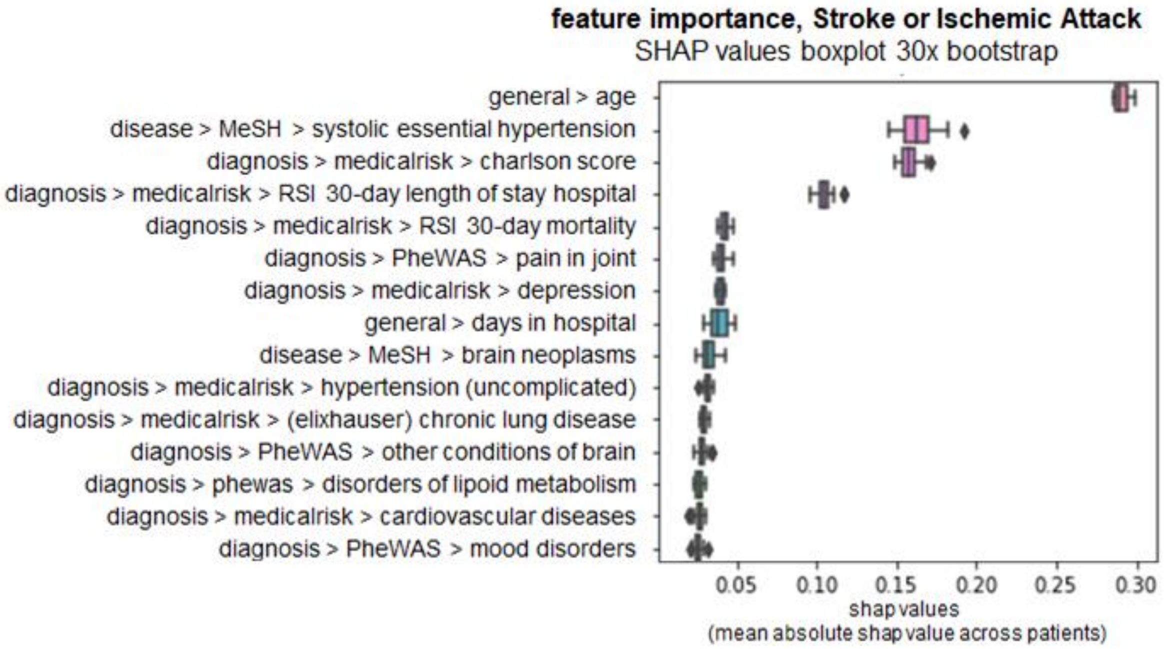


**figure S 6:** SHAP boxplot feature importance Stroke or Ischemic Attack

##### 5.2.1 Marginal dependency plots for top 5 features

The following plots show marginal influences of the top 5 features (according to the mean absolute SHAP value, see before) on predicted comorbidity risks.

- x-axis denotes feature values
- y-axis denotes the SHAP value (higher value = higher influence on predicted comorbidity risk (hazard rate) compared to average patient)

##### *5.2.1.1 Anxiety*

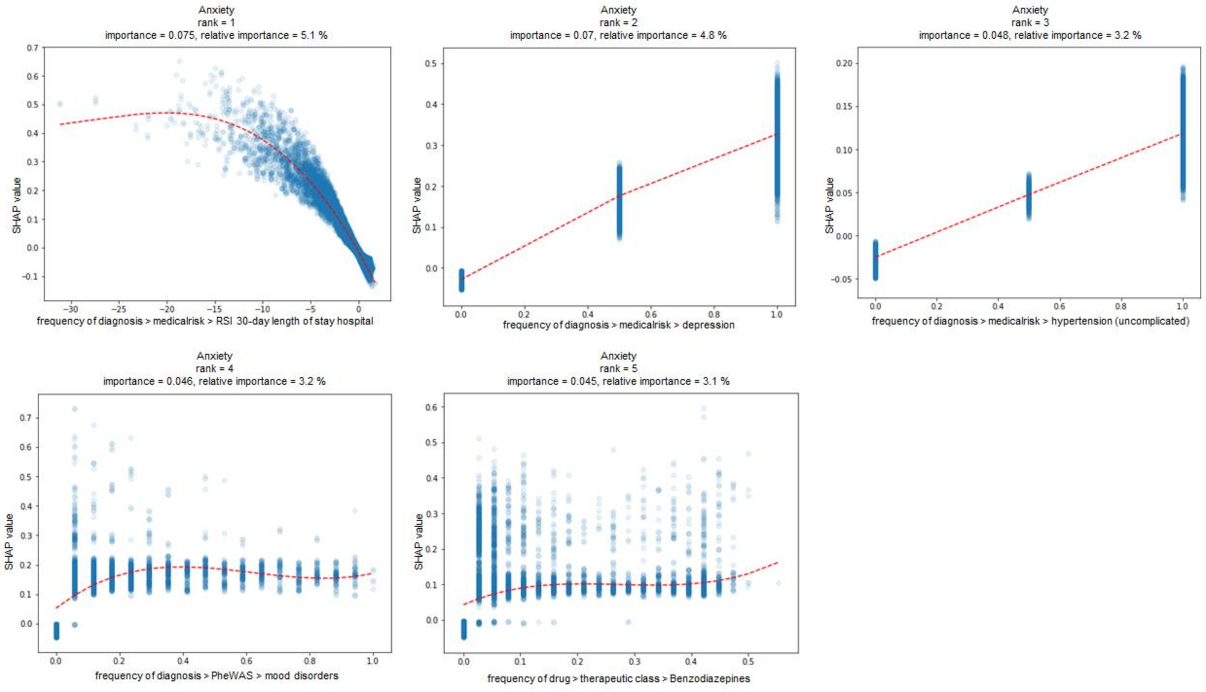


**Figure S 7:** SHAP marginal dependency plot Anxiety

##### *5.2.1.2 Bipolar, Schizophrenia*

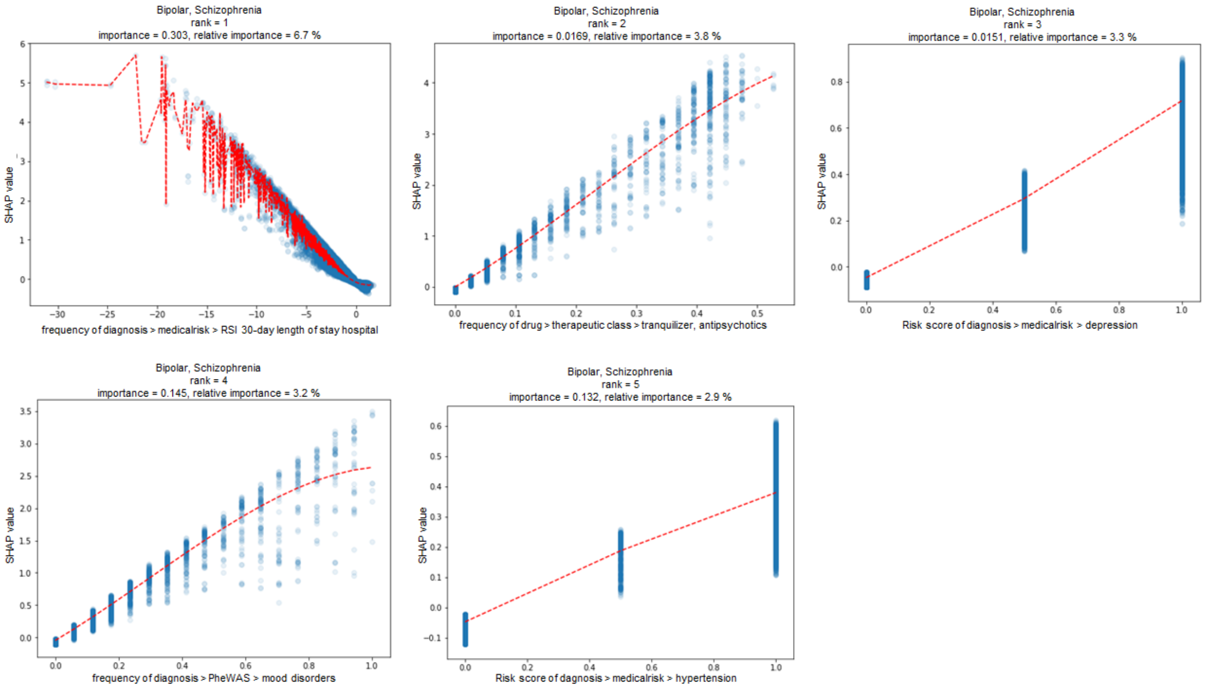


**Figure S 8:** SHAP marginal dependency plot Bipolar, Schizophrenia

##### 5.2.1.3 *Diabetes type 2*

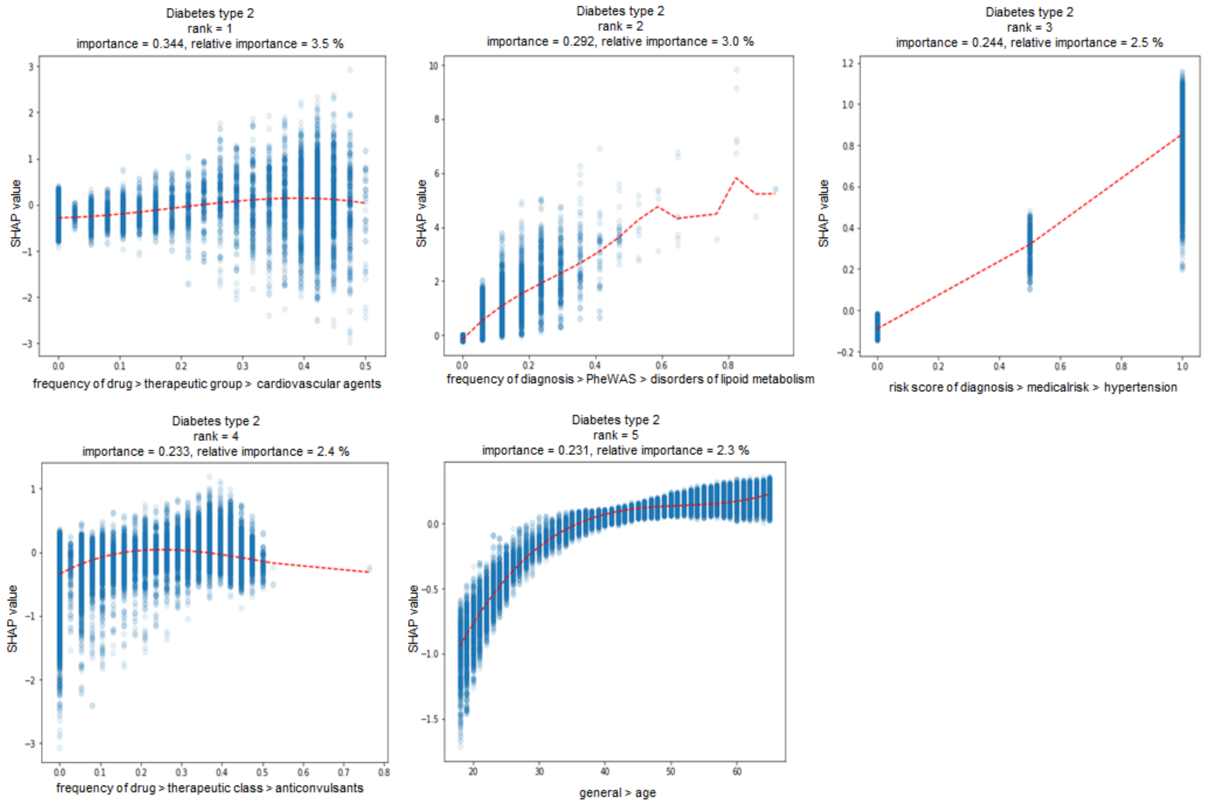


**Figure S 9:** SHAP marginal dependency plot Diabetes type 2

##### 5.2.1.4 *Migraine*

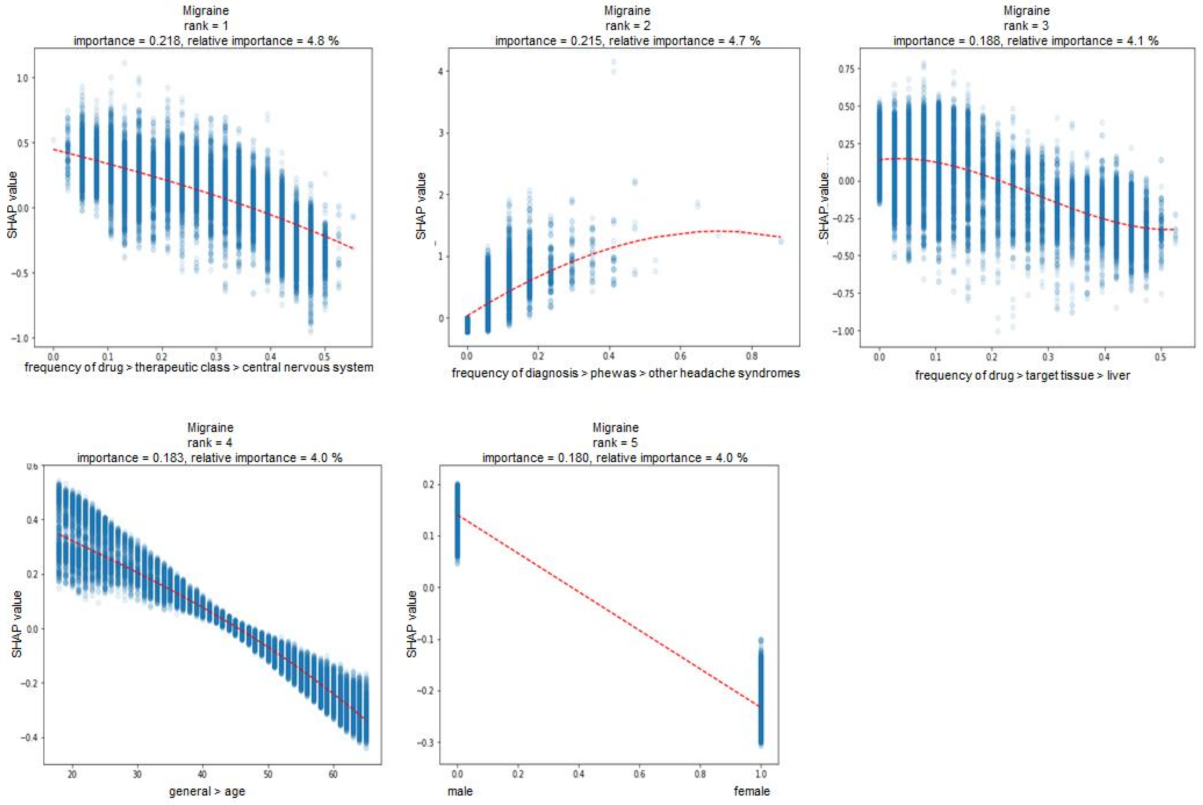


**Figure S 10:** SHAP marginal dependency plot Migraine

##### 5.2.1.5 *Overweight, Obesity*

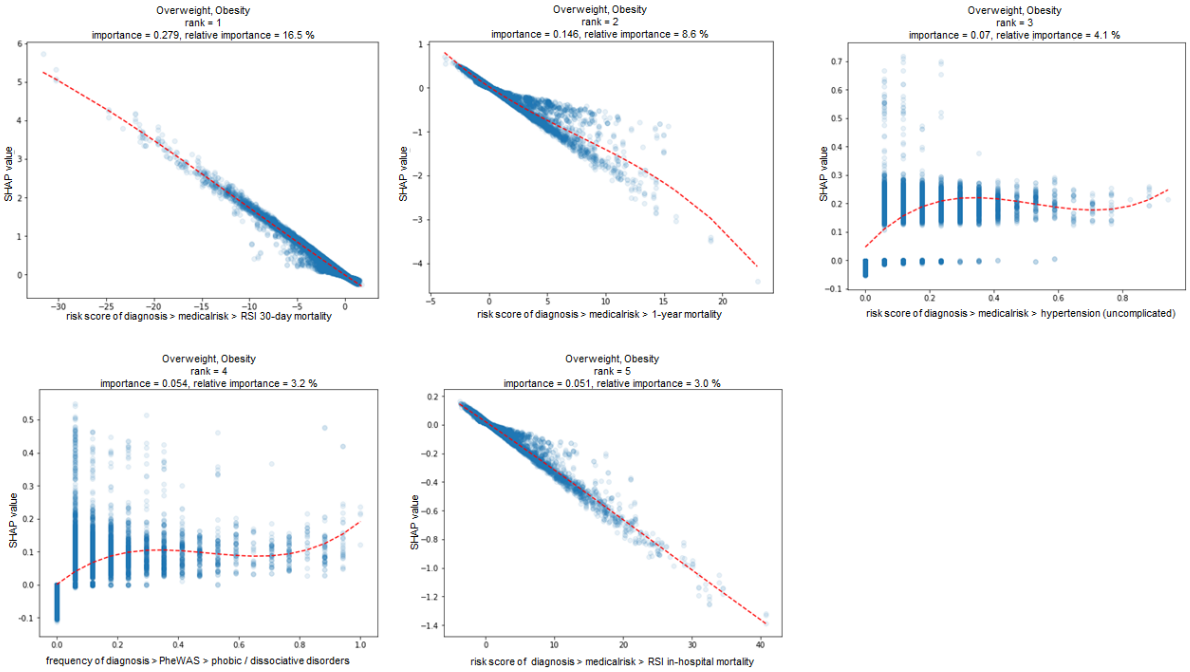


**figure S 11:** SHAP marginal dependency plot Overweight, Obesity

##### 5.2.1.6 *Stroke, Ischemic Attack*

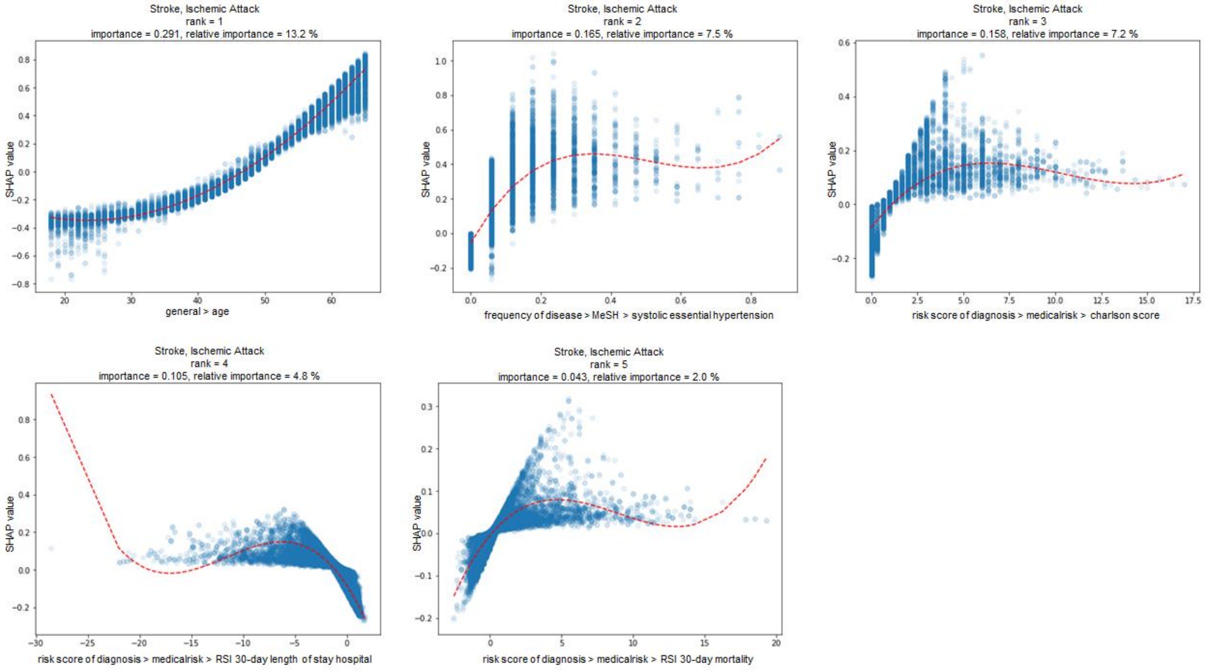


**figure S 12:** SHAP marginal dependency plot Stroke, Ischemic Attack
